# Limited genomic reconstruction of SARS-CoV-2 transmission history within local epidemiological clusters

**DOI:** 10.1101/2021.08.08.21261673

**Authors:** Pilar Gallego-García, Nair Varela, Nuria Estévez-Gómez, Loretta De Chiara, Iria Fernández-Silva, Diana Valverde, Nicolae Sapoval, Todd Treangen, Benito Regueiro, Jorge Julio Cabrera-Alvargonzález, Víctor del Campo, Sonia Pérez, David Posada

## Abstract

A detailed understanding of how and when SARS-CoV-2 transmission occurs is crucial for designing effective prevention measures. Other than contact tracing, genome sequencing provides information to help infer who infected whom. However, the effectiveness of the genomic approach in this context depends on both (high enough) mutation and (low enough) transmission rates. Today, the level of resolution that we can obtain when describing SARS-CoV-2 outbreaks using just genomic information alone remains unclear. In order to answer this question, we sequenced 49 SARS-CoV-2 patient samples from ten local clusters for which partial epidemiological information was available, and inferred transmission history using genomic variants. Importantly, we obtained high-quality genomic data, sequencing each sample twice and using unique barcodes to exclude cross-sample contamination. Phylogenetic and cluster analyses showed that consensus genomes were generally sufficient to discriminate among independent transmission clusters. However, levels of intrahost variation were low, which prevented in most cases the unambiguous identification of direct transmission events. After filtering out recurrent variants across clusters, the genomic data were generally compatible with the epidemiological information but did not support specific transmission events over possible alternatives. We estimated the effective transmission bottleneck size to be 1-2 viral particles for sample pairs whose donor-recipient relationship was likely. Our analyses suggest that intrahost genomic variation in SARS-CoV-2 might be generally limited and that homoplasy and recurrent errors complicate identifying shared intrahost variants. Reliable reconstruction of direct SARS-CoV-2 transmission based solely on genomic data seems hindered by a slow mutation rate, potential convergent events, and technical artifacts. Detailed contact tracing seems essential in most cases to study SARS-CoV-2 transmission at high resolution.

## Introduction

In recent years, genomic epidemiology has revealed itself as a powerful tool for tracking viral outbreaks (Grubaugh, Ladner, et al. 2019). Particularly for diseases with a high proportion of asymptomatic infections like COVID-19, the use of genomic information might be especially relevant to understand their dissemination. Several methods have been developed to reconstruct infectious disease outbreaks using genomic information (e.g., Didelot et al. 2014; Jombart et al. 2014; Worby, Chang, et al. 2014; Hall et al. 2015; Hall et al. 2016; Lumby et al. 2018; Didelot et al. 2021). However, these strategies rely on pathogen genomes mutating rapidly between infected individuals (Campbell et al. 2018). Severe acute respiratory syndrome coronavirus 2 (SARS-CoV-2), responsible for the COVID-19 pandemic, has spread globally in a very short time. SARS-CoV-2 has a mutation rate in the order of 1 × 10^−3^ mutations per site per year (van Dorp, Richard, et al. 2020; Koyama et al. 2020). For MERS-CoV-2, in principle with a similar mutation rate, the prediction is that in most cases, the consensus sequences sampled from a transmission pair (donor and receptor) will be identical, precluding a complete reconstruction of the outbreak (Campbell et al. 2018). As a counterpart, for SARS-CoV-1, with a mutation rate four times higher, we expect to see several mutations between transmission pairs, which considerably augments the power to resolve transmission history (Campbell et al. 2018).

These considerations are based on consensus sequences that represent the dominant viral lineage within a host. However, pathogens with high rates of evolution, such as RNA viruses, accumulate new mutations more or less rapidly as they replicate within the individuals they infect, generating intrahost genomic variation. The generation of this genomic diversity enables viral populations to evade host immune responses (Hensley et al. 2009; Henn et al. 2012; Parameswaran et al. 2017), alter disease severity (Vignuzzi et al. 2006), and adapt to changing environments (Stapleford et al. 2014; Stern et al. 2017). Notably, the study of the shared intrahost genomic variation among individuals can be critical for identifying contagion events and transmission clusters (Didelot et al. 2014; Worby et al. 2014; Park et al. 2015; Worby et al. 2017). Moreover, it also allows for estimating the size of the founding pathogen population transmitted from the donor to the recipient host (i.e., the transmission bottleneck size) (Frise et al. 2016; Sobel Leonard et al. 2017; Sobel Leonard et al. 2019). Several studies have already shown that intrahost genomic variation can be detected in most SARS-CoV-2 infections, generally at low levels, but with some variation among individuals (Kuipers et al. 2020; Seemann et al. 2020; Shen et al. 2020; Tonkin-Hill et al. 2020; Wölfel et al. 2020; Butler et al. 2021; Lythgoe et al. 2021; Valesano et al. 2021; Y. Wang et al. 2021). Most SARS-CoV-2 intrahost mutations appear at low frequencies, often less than 5%, are primarily under purifying selection, and display particular biochemical signatures (Tonkin-Hill et al. 2020; Graudenzi et al. 2021; Sapoval et al. 2021; Y. Wang et al. 2021).

A key question is whether SARS-CoV-2 intrahost variation can be transmitted during contagion. The answer is not straightforward, as shared intrahost variants among unrelated individuals can also result from convergent evolution or mutational hotspots (Tonkin-Hill et al. 2020; Valesano et al. 2021). So far, a few studies have used shared genomic variants between putative donor-receptor pairs to infer (narrow) transmission bottlenecks, of 1-10 virions, in SARS-CoV-2 (Li et al. 2021; Lythgoe et al. 2021; San et al. 2021; D. Wang et al. 2021). Limited genomic diversity can prevent the reconstruction of disease outbreaks (Campbell et al. 2018). While distinct SARS-CoV-2 transmission clusters might be identified using consensus sequences (Letizia et al. 2020; Popa et al. 2020; Seemann et al. 2020), its moderate mutation rate and rapid transmission might prevent the detailed reconstruction of the transmission events within these clusters (Tonkin-Hill et al. 2020). Leveraging intrahost variation, San et al. (2021) studied two nosocomial SARS-CoV-2 outbreaks, showing that potential donor-recipient pairs are supported in some cases but not in others by shared intrahost variants.

All in all, it is not clear whether the observed levels of inter and intrahost variation in SARS-CoV-2 and the apparently small size of the transmission bottleneck could limit our capability to reconstruct local SARS-CoV-2 outbreaks in detail using only genomic information. Intrahost mutations, typically at very low frequencies, are sensitive to methodological artifacts like sequencing errors (De Maio et al. 2020a; Turakhia et al. 2020; Kubik et al. 2021) and cross-sample contamination, and the occurrence of mutational hotspots can confound the identification of transmission events. Here, we wanted to assess our ability to reconstruct putative transmission chains and to infer reliable transmission bottleneck sizes in SARS-CoV-2. For this, we obtained high-quality genomic data from ten independent epidemiological clusters, with two replicates per sample and with unique oligonucleotide spike-ins to detect potential contamination, leveraging both interhost and intrahost variants and *ad hoc* phylogenetic techniques. Our results confirm the low levels of intrahost variability and the small transmission bottleneck of SARS-CoV-2, suggesting that genomic data alone might not be sufficient to fully resolve direct SARS-CoV-2 transmissions, revealing the need for additional sources of information like detailed contact tracing.

## Material and methods

### Sample collection

According to the epidemiological records, we identified 49 patients infected with SARS-CoV-2 conforming ten independent transmission clusters originated in nursing homes, family households, and birthday parties from the same city (**Figure 1**; **Table S1**). After that, we recovered the corresponding diagnostic nasopharyngeal exudates collected. This study was conducted under the approval of the Galician Drug Research Ethics Committee (CEIm-G code 2020-301).

**Figure 1.**
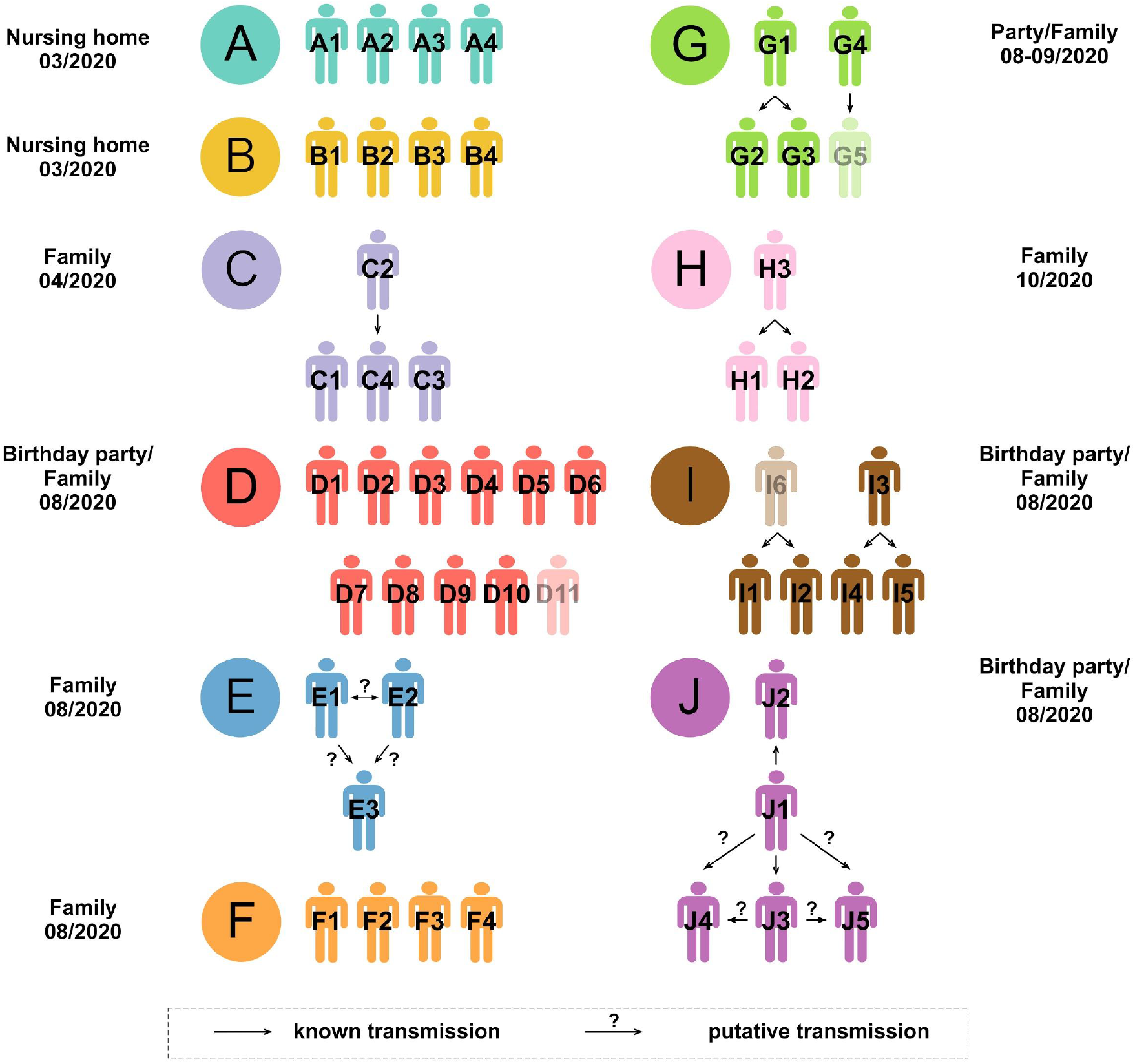
Transmission clusters and epidemiological information. Black arrows indicate “known” transmission events identified in the epidemiological records. Question marks highlight potential alternatives. Samples from patients in faded color failed at sequencing.

### Epidemiological information

Cluster A and B belong to two different nursing homes, and in both cases, the primary case could not be established with confidence (**Figure 1**). Cluster C is a family in which there was a probable transmission from C2 to C4. Cluster D is a large family spanning four different households. D1 came from another Spanish city and likely started the D transmission at a birthday party. Cluster E is a family in which brothers E1 and E2 were infected abroad before infecting their parent, E3. Cluster F is another family that was likely infected by an unsampled case from another city. Cluster G originates in two individuals (G1 and G4) that attended the same event and afterward infected their respective families, G1 to G2 and G3, and G4 to G5 (G5 failed at sequencing). Cluster H is another family in which H3 likely infected H1 and H2. Cluster I starts with two children (I6 and I3; I6 failed at sequencing) that got infected at the same birthday party before infecting their families, I6 to I1 and I2, and I3 to I4 and I5. Cluster J is a family in which J1 infected partner J2 and child J3. After that, either J1 or J3 infected J4 and J5.

### RNA extraction

Following the manufacturer’s recommendations, we extracted the viral RNA from the nasopharyngeal exudates using the MagNA Pure 24 Total NA Isolation kit (Roche Diagnostics, Basel, Switzerland). Different team members processed each RNA sample independently to obtain two technical replicates for each patient sample, from retrotranscription to library construction.

### Viral load measurement

We measured SARS-CoV-2 genome copy concentration for each sample by real-time RT-PCR of the E gene with the Sarbecovirus E-gene ModularDx (TIB Molbiol, Berlin, Germany) kit in a LightCycler® z480 System (Roche Molecular Systems Inc, Meylan, France). Viral load was estimated using linear regression (R^2^ >0.99) from the standard curve generated with the Ct values obtained for serial dilutions (log) of RNA standards with known viral RNA genome equivalents / µL (Vogels et al. 2020).

### cDNA synthesis and multiplex amplification

We followed the ARTIC sequencing protocol (v.3) (Quick et al. 2017), a multiplex PCR-based target enrichment that produces 400 bp amplicons that span the SARS-CoV-2 genome, with slight modifications. First, we retrotranscribed the RNA samples to cDNA using the SuperScript IV reverse transcriptase (Invitrogen, MA, USA), starting with 10 µL of RNA. Then we ran 30 PCR cycles for all the samples, independently of the Ct value, using the ARTIC primer Pool1 and Pool2 (IDT, CA, USA) and the Q5 Hot Start DNA polymerase (New England Biolabs, MA, USA). Next, we mixed the corresponding PCR products from each sample before cleaning (1.2:1 ratio beads to sample). We eluted the clean PCR products with 35 µL NFW, recovered 33 µL and performed quantification with the Qubit 3.0 using the dsDNA HS or BR kit (Thermo Fisher Scientific, MA, USA) and checked amplicon size with the 2200 TapeStation D1000 kit (Agilent Technologies, CA, USA).

### Addition of individual barcodes

We added 1 µL of an X-mer single-stranded oligonucleotide with a unique barcode sequence at 38 fM to each retrotranscription reaction to detect potential sample cross-contamination. To prepare these barcode spike-ins, we used as a template the alcohol dehydrogenase 1 (*adh1*) mRNA (XM_008650471.2) from *Zea mays*, as described in the PrimalSeq v.4.0 protocol (Matteson et al. 2020). After a cleanup step (2:1 ratio beads to sample), we recovered a final volume of 22 µL and performed QC (Qubit 3.0 and 2200 TapeStation). We added F and R primers with the same barcode sequence at the same concentration as the ARTIC primer pools to amplify the barcodes in the multiplex PCR reactions.

### Library construction and genome sequencing

We built 98 whole-genome sequencing libraries employing the DNA Prep (M) Tagmentation kit (Illumina, CA, USA) using ¼ of the recommended volume, with approximately 125 ng of input DNA. Finally, we checked the size of the libraries and quantified them as described above. We sequenced the 98 libraries in two high-output (7.5 Gb) runs (60 and 38 samples, respectively) on an Illumina MiniSeq (PE150 reads) at the sequencing facility of the University of Vigo.

### Detection of potential cross-sample contamination

To assess the level of cross-sample contamination, we quantified the specific maize barcode content in each fastq file. For this, we aligned the raw reads against the *Zea mays adh1* sequence using BWA-mem (Li 2013) with default settings and counted the number of reverse and forward barcodes with cutadapt (v.2.10) (Martin 2011), with a minimum overlap of 15 and a maximum error rate of 0.1.

### Variant calling and consensus sequences

We assessed the quality of the fastq files using FastQC (Andrews 2010). Then we aligned the reads to the reference MN908947.3 from Wuhan using BWA-mem (Li 2013) and trimmed them with iVar (Grubaugh, Gangavarapu, et al. 2019). We evaluated the quality of the aligned trimmed reads using Picard v2.21.8 (http://broadinstitute.github.io/picard). We used SAMtools depth v1.10 [Citation error] to calculate the sequencing coverage along the genome for each replicate. We only kept samples for which ten or more reads covered more than 75% of the viral genome in the two replicates and with less than 2.5% missing bases on the consensus sequence.

We used iVar (Grubaugh, Gangavarapu, et al. 2019) to identify single nucleotide changes and indels, with a minimum base quality threshold of 20 and a minimum read depth of 10. The calls obtained were confirmed with LoFreq (Wilm et al. 2012). We only retained variants that appeared in both replicates with a minimum overall variant allele frequency (VAF) of 2%. Based on their frequency, we divided the genomic changes detected into *fixed* (VAF ≥ 0.98; to account for potential sequencing errors) and *intrahost* variants (0.02 ≤ VAF < 0.98). We masked and removed from further analyses positions containing complex variants (i.e., nucleotide changes plus indels) or those deemed as homoplasic (De Maio et al. 2020b), including the sites immediately before and after.

To build a consensus sequence for each sample, we merged the reads from the two replicates with SAMtools *mpileup* and fed them to iVar *consensus* with a minimum VAF threshold of 0.5. We assigned the consensus sequences to a SARS-CoV-2 clade with Nextclade (https://clades.nextstrain.org) and to a SARS-CoV-2 PANGO lineage (Rambaut et al. 2020) with Pangolin (O’Toole et al. 2021).

### Delimitation of epidemiological clusters

The simplest method for delimiting epidemiological clusters using genomic data alone is estimating a phylogenetic tree using the consensus sequences. For this, we aligned the consensus sequences with the reference using MAFFT v.7 (Katoh and Standley 2013) (*mafft --maxiterate 500 <input>*) and ran IQ-TREE (v.2.0.6) (Nguyen et al. 2015) (*iqtree2 -T AUTO -s <aligment*.*fa> -m TEST -b 1000 -o MN908947*.*3)* with the best-fit nucleotide substitution model and 1,000 bootstrap replicates. We also built a timetree based on the output tree of IQ-TREE and the dates of the samples using TreeTime (v.0.8.1) (Sagulenko et al. 2018) (*treetime --aln <alignment*.*fa> --tree <treename> --dates <dates*.*csv>*). In addition, we tried six heuristics developed explicitly for the reconstruction of epidemiological clusters described in Worby et al. (2017). The weighted distance tree and the minimum distance tree use the genetic distances among consensus sequences.

On the other hand, the weighted and maximum variant tree strategies rely exclusively on shared intrahost variants. Finally, the hybrid weighted tree and maximum tree procedures use intrahost variants and consensus genetic distances. Furthermore, we also estimated transmission clusters using the Transcluster algorithm (Stimson et al. 2019), assuming a mutation rate of 1 × 10^−3^ mutations/site/year. We explored four values for the transmission rate (10, 25, 50, 100 transmissions per year) and six for the transmission cutoff (1–6 transmission events).

### Inference of transmission history

Within each cluster, we tried several approaches to estimate which individuals transmitted the virus and in which direction, that is, to learn who infected whom. First, we explored the Worby et al. heuristics, which assume that the donor/s for each sample has the most similar sequence or more shared intrahost variants. In addition, we implemented a simple approach that leverages the intrahost variation along a minimum spanning tree (MST). First, we computed Euclidean pairwise distances among all individuals within a cluster, with the *rdist* R package (https://github.com/blasern/rdist), using the VAF distributions. Afterward, we built the MSTs based on those distances with the function *mst* from the *ape* R package (Paradis and Schliep 2019). Then, assuming a single source for each cluster, we inferred the transmission direction that minimized the generation of novel variants in the receptor, meaning that in a pair of individuals, the donor should be the one with a higher number of private mutations. Finally, we also explored TransPhylo (Didelot et al. 2017), using the dated phylogeny obtained with TreeTime. We ran the algorithm for 150,000 MCMC iterations and assumed a Gamma distribution for the generation time with shape 1 and scale 0.01917 (Perera et al. 2021).

### Estimation of the transmission bottleneck size

To estimate the transmission bottleneck size of SARS-CoV-2 (i.e., the size of the viral population transferred from the donor to the recipient host), we used the beta-binomial method of Sobel Leonard et al. (2017). This method assumes that the intrahost variants detected did not arise *de novo* in different patients. This calculation includes only intrahost donor variants shared with the recipient (note that they can be fixed in the recipient but not in the donor). We identified putative donor-recipient pairs according to the available epidemiological information (**Figure 1**). We lacked epidemiological information for clusters D and F, and we identified possible transmission pairs according to the genomic data (see Results). For the estimation of the transmission bottleneck size, we used the R code at https://github.com/weissmanlab/BB_bottleneck (version of March 24, 2020), under the *approximate* model (given that the sequencing depth per sample was very high, around 6,000X) and setting the maximum bottleneck size to an arbitrarily large value of 600, and the VAF cutoff to 0.02.

### Assessment of selective pressures

The ratio of non-synonymous changes per non-synonymous site (*dN*) to the number of synonymous substitutions per synonymous site (*dS*) is one of the most popular statistics for detecting selective pressures at the molecular level. We estimated the *dN/dS* ratio for each sample using the dNdScv package (Martincorena et al. 2017), recently adapted for its application to SARS-CoV-2 (Tonkin-Hill et al. 2020). We used the default substitution model with 192 rate parameters.

## Results

### Viral load and sequencing

Twenty-seven out of the 49 samples had a viral load above 10^3^ copies / µL (**Table S1**). Sequencing coverage and breadth were high (mean depth ± sd: 6316.71 ± 2336.99; breadth: 0.999) (**Table S2**), except for three samples (D11, G5, and I6, all with a Ct > 32 for gene E), that we excluded from further analyses. We did not detect appreciable cross-contamination between samples (**Table S2**).

### Inter and intrahost variation

Most variants were fixed (**Figure 2, Figure S1**). The number of differences among consensus sequences was, on average, 2.12, 2.28, and 7.57 variants, within early clusters (A-C), late clusters (D-J), and among early and late clusters (see also **Table S3**). We observed on average 19.76 variants per sample, of which 8.17 were intrahost (**Table S4**). Both fixed and intrahost variants were shared among samples at different VAFs. Several intrahost variants appeared recurrently in multiple samples, often corresponding to indels at low frequency (**Table S4**). These recurrent variants may correspond to potential sequencing errors and mutational hotspots, which might confound our analyses. Therefore, we decided to filter out intrahost variants present in more than one cluster. After filtering, there were 2.13 intrahost variants per sample on average, with a maximum of 11 (**Table S4**). Before and after filtering, the number of intrahost variants detected per sample was unrelated to sequencing depth, Ct values, or viral load (**Figure S2**). Furthermore, VAFs between sample replicates were significantly correlated (Pearson correlation coefficient = 0.99, p-value = 5.6 e-113) (**Figure S3**). All samples were assigned to two related clades/lineages (20A/B.1 and 20E(EU1)/B.1.177), which was not particularly surprising as these were the dominant lineages in the area at the time of sampling.

**Figure 2.**
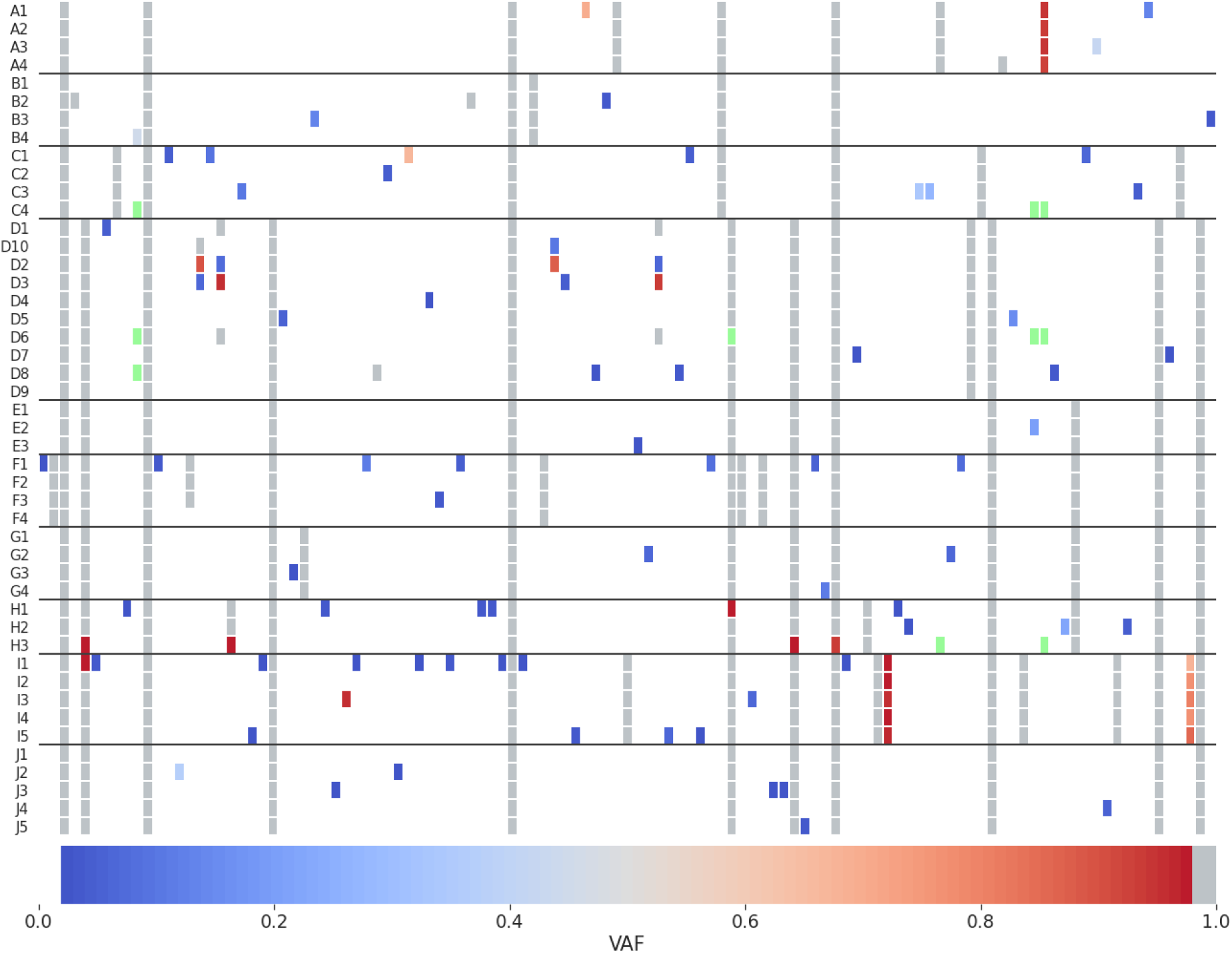
Variant allele frequencies (VAF) per sample. VAFs were calculated after filtering recurrent variants. Fixed mutations (VAF ≥ 0.98) are in gray, fixed reference alleles (VAF < 0.02) are in white, and positions with missing data (depth below 20) are in light green.

### Delimitation of transmission clusters

The maximum likelihood (ML) trees obtained with the consensus sequences showed the epidemiological clusters as distinct groups, mostly monophyletic and with relatively high bootstrap support (**Figure 3**). However, standard phylogenetic approaches do not explicitly inform about the number of clusters or the assignment of the different individuals to clusters. In the absence of additional epidemiological information (i.e., colors in the tree), it is up to the researcher to decide where to “cut”. Remarkably, adding the temporal information with TreeTime improves the phylogenetic resolution of the clusters, which become all monophyletic (**Figure 3B**).

**Figure 3.**
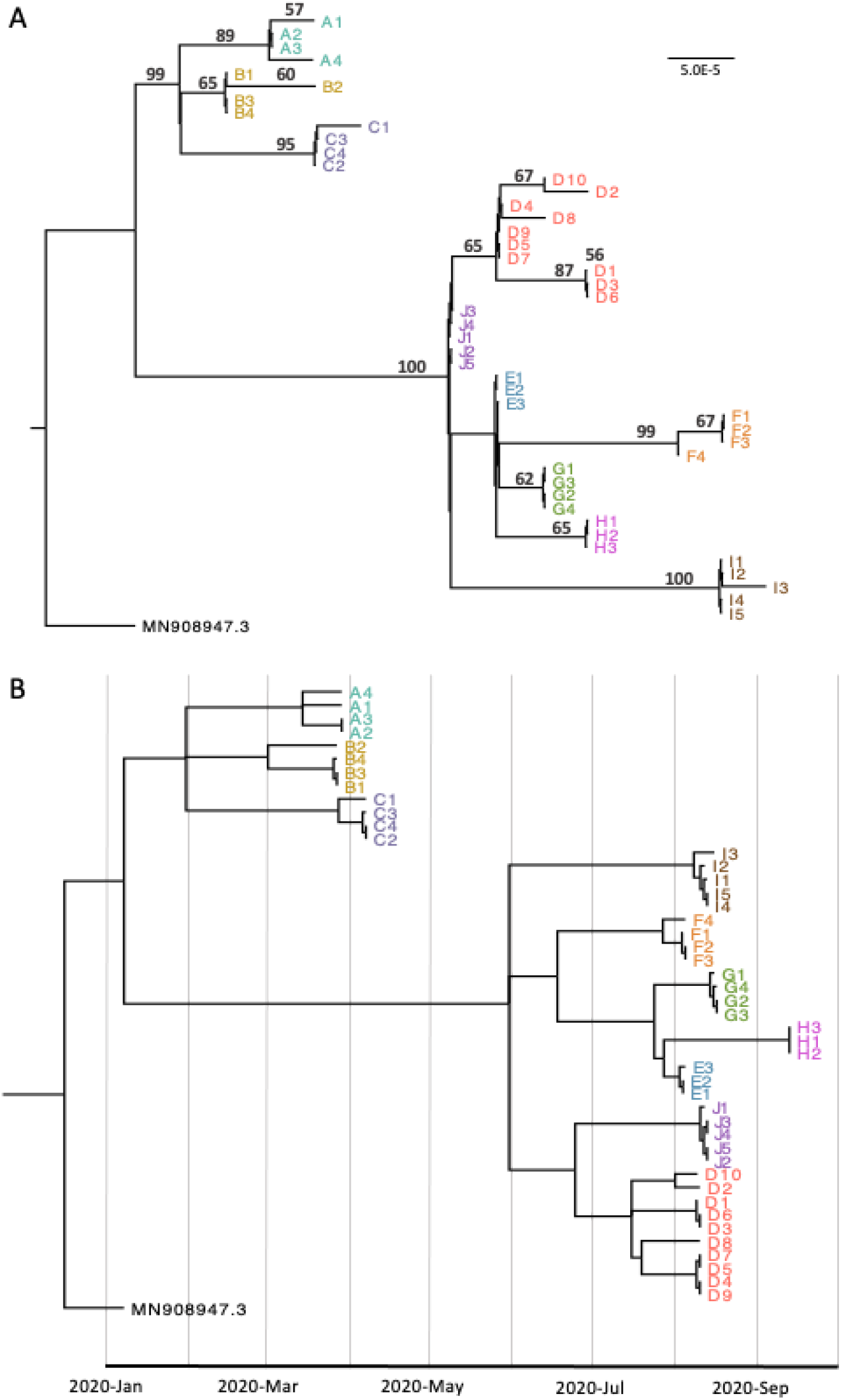
Consensus-sequence phylogenetic trees. (A) Maximum likelihood phylogenetic tree inferred with IQ-TREE. Numbers above branches are bootstraps values (%). Only bootstrap values above 50 are shown. (B) Time-scaled ML tree inferred with TreeTime (using the dates of extraction).

The weighted distance tree and the minimum distance tree, which also use consensus sequences but can explicitly delimit clusters, were identical and highly congruent with the epidemiological information (**Figure 4A**). In this case, the only “error” was that cluster D was divided into two, although we might expect it because D1, D3, and D6 share two consensus mutations that the rest of the D individuals do not present. Indeed, cluster D is large and phylogenetically diverse, and we might not have sampled all the infected individuals in this transmission chain. The weighted variant and maximum variant trees, based exclusively on intra-host variants, were also identical and produced a very complex network in which all individuals seemed related to each other (**Figure 4B**). After removing the recurrent intra-host variants common to multiple individuals and clusters (taking advantage of the epidemiological information), these methods identified three clusters primarily compatible with the epidemiological information, plus 33 unconnected individuals (**Figure 4C**). Cluster A was perfectly delimited, while cluster I formed a group with a sample from cluster H. The only other three clusterized samples were from cluster D (again D1, D3, and D6). The hybrid transmission methods, which use the connections established by the weighted variant and maximum variant trees and incorporate consensus information for those samples without a donor or recipient, did not result in any noticeable improvement compared to methods based on consensus sequences (data not shown). Finally, the transmission-based clustering method in Transcluster was able to identify some of the epidemiological clusters but not all (**Figure 4D**). In this case, congruence with the epidemiological data was maximal after setting a transmission rate of 25 and a transmission cutoff of 1.

**Figure 4.**
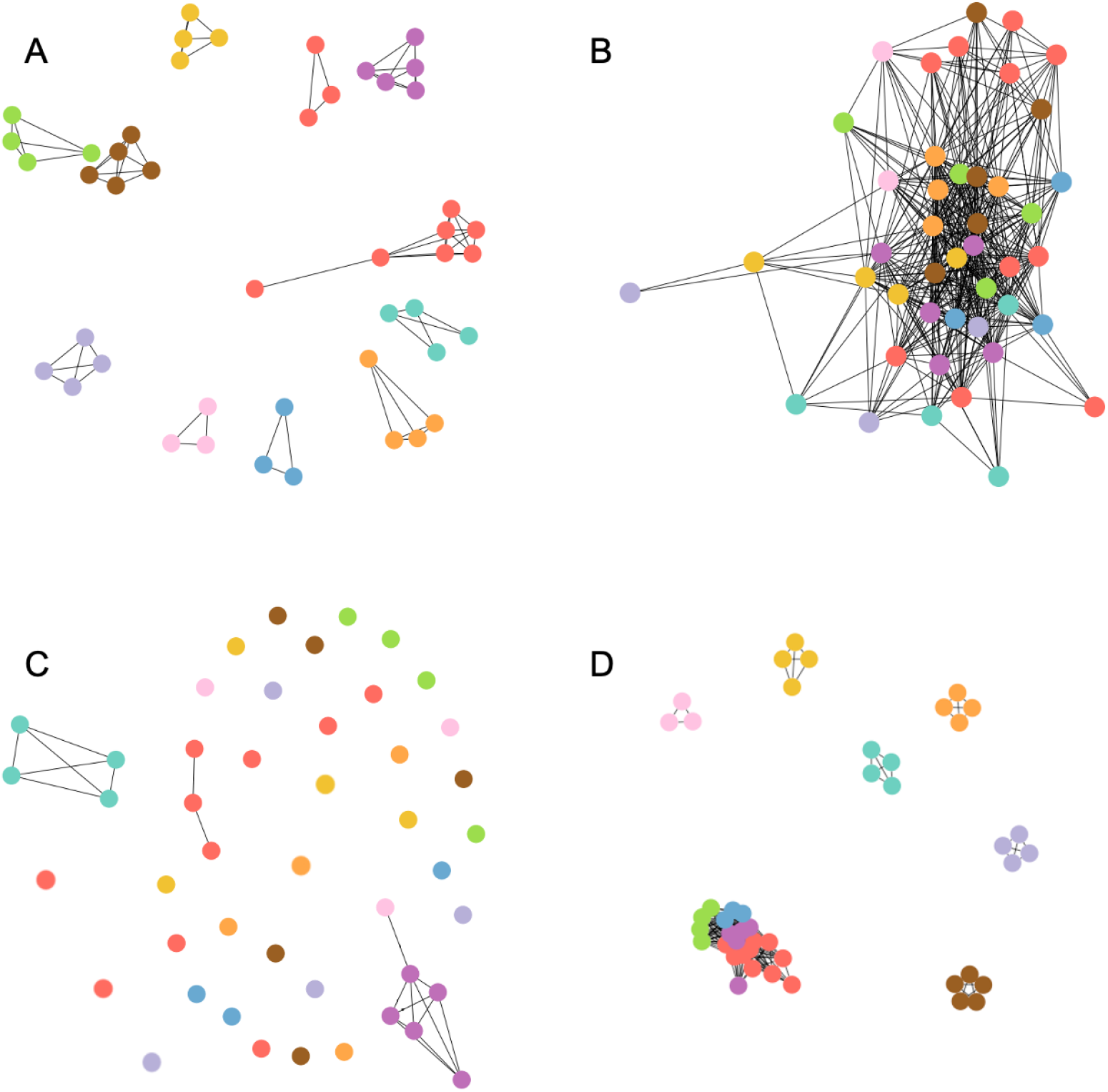
Clustering approaches. (A) Weighted distance/minimum distance tree. (B) Weighted variant/maximum variant tree after standard masking. (C) Weighted variant/maximum variant tree after removing recurrent low-frequency variants (D) Transcluster transmission network (transmission rate = 25, transmission cutoff = 1).

### Inference of transmission history

#### Transmission in nursing homes

For clusters A and B, we had no epidemiological information other than the corresponding nursing home. In cluster A (**Figure 5**), the four samples share what seems to be an intrahost variant (27695-TCTTA). However, given its high VAF (0.93-0.95) and the fact that the individuals do not share other variants, this deletion may be a fixed variant, where sequencing or calling errors prevented its identification in all reads. In any case, the genetic data does not help identify the different transmission events in this cluster with confidence. In cluster B, no shared variation was apparent. B2 has two private fixed variants, suggesting it was infected later than the other cluster members or from a different source. Again, it was not possible to infer the transmission network for this cluster.

**Figure 5.**
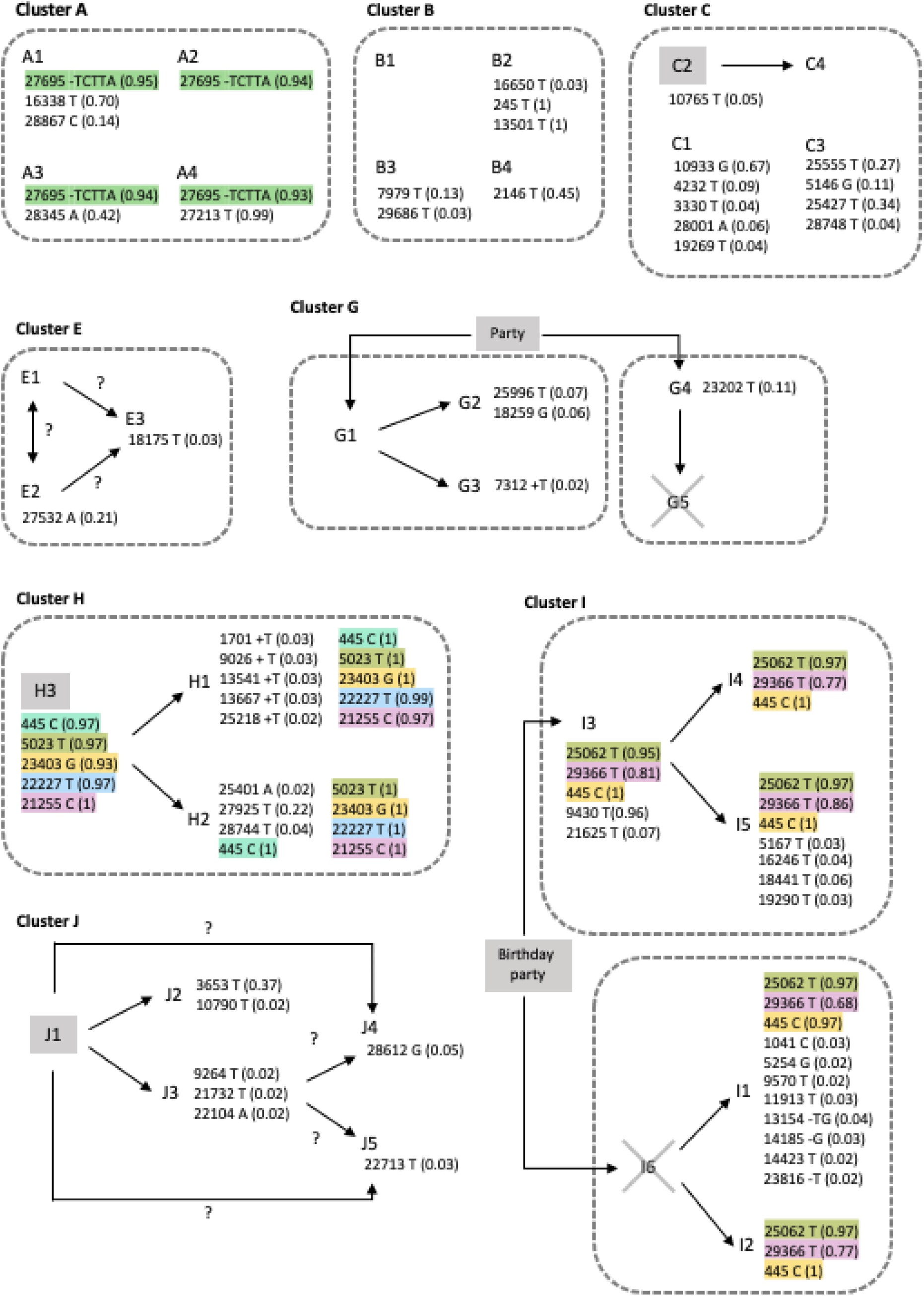
Variant sharing within clusters. Gray boxes indicate index cases or originating events. Gray dashed lines delimit households (nursery homes for clusters A and B). Shared variants are highlighted with the same color. Fixed variants (VAF ≥ 0.98) common to all members of a cluster are not shown. Crossed samples could not be sequenced.

#### Transmission in clusters with partial contract trace information

We had partial contact trace information for clusters C, E, G, H, I, and J. However, the lack of shared intrahost variants prevented a detailed reconstruction of their transmission history in most cases (**Figure 5**). The epidemiological record suggests a transmission from C2, the index case, to C4 in cluster C. This event is compatible with the genetic data, as C2 has a single intrahost variant at low frequency (0.05), which could have been lost during transmission to C4, which has no intrahost variants. Private variants with low VAFs in C1 and C3 could have arisen de novo within each individual after transmission, but three of them with higher VAFs (0.27, 0.34, and 0.67) are more difficult to explain in the same way. In cluster E, the genetic information cannot resolve whether E1 or E2 infected E3. In cluster G, we did not observe shared intrahost variants. In contrast, the distribution of the private variants is compatible with the epidemiological information, and it does not help resolve further the transmission history.

In cluster H, the three samples share five fixed (or almost fixed) variants. The index case (H3) does not seem to have intrahost variants, contrary to H1 and H2. However, the quality of the sequencing data in the case of H3 is well below average, so it is possible that low-frequency variants were overlooked in this sample. All five members of cluster I share three variants with high VAF –or fixed in several cases. Variants 445C and 25062 could be genuinely fixed in all samples, including cases where the apparent VAF is 0.95-0.97. The distribution of variant 29366T is remarkable, as it appears in all cases with a VAF of 0.68-0.86. Another salient observation is that I3, one of the index cases, has a well-supported variant (9430T) with a VAF of 0.96 that does not appear in the other samples from this cluster. Cluster J lacked shared intrahost variants, so the genetic data neither confirmed nor invalidated the contact tracing information.

#### Inferring transmission in the absence of contact trace information

##### Ad hoc approaches

We did not have detailed information about contacts in clusters D and F, so we tried to identify transmission events considering just the genomic data (**Figure 6**). In cluster D, the transmission started at a birthday party where the index case was D1. D1 shares two variants with D3 and D6 (4543T and 18431T), both fixed in D1 and D6 and close to fixation in D3 (0.96 and 0.94, respectively). Therefore, we hypothesize that D1 → D3 and D1 → D6, but alternatively D3 → D6, could be transmission pairs. These two variants also appear in D2 but at a very low VAF (0.07 and 0.06, respectively). D3 and D2 further share 4142 A, but this variant has a low VAF in D3 (0.07) and a high VAF (0.89) in D2. Furthermore, D2 has 15857T at high VAF (0.88). Given that we assumed that D1 infected D3, we considered that D3 could have infected D2. However, the explanation for the observed VAF patterns might imply recombination and *de novo* mutation. Finally, D10 shares with D2 variants 4142A and 15857T at high frequency, so we also considered D2 → D10 another likely transmission pair. In cluster F, where we do not have an index case, F1 and F3 share a fixed variant (3737 T). Given that F1 has seven private variants at low frequency, but F3 only two, F1 might be the pair’s donor because it seems easier to lose these variants during the F1 → F3 bottleneck than to arise d*e novo* in F1 after an F3 → F1 transmission. F2 also has 3737 T fixed. Following the same logic, F2 could have been infected by F1, but also by F3.

**Figure 6.**
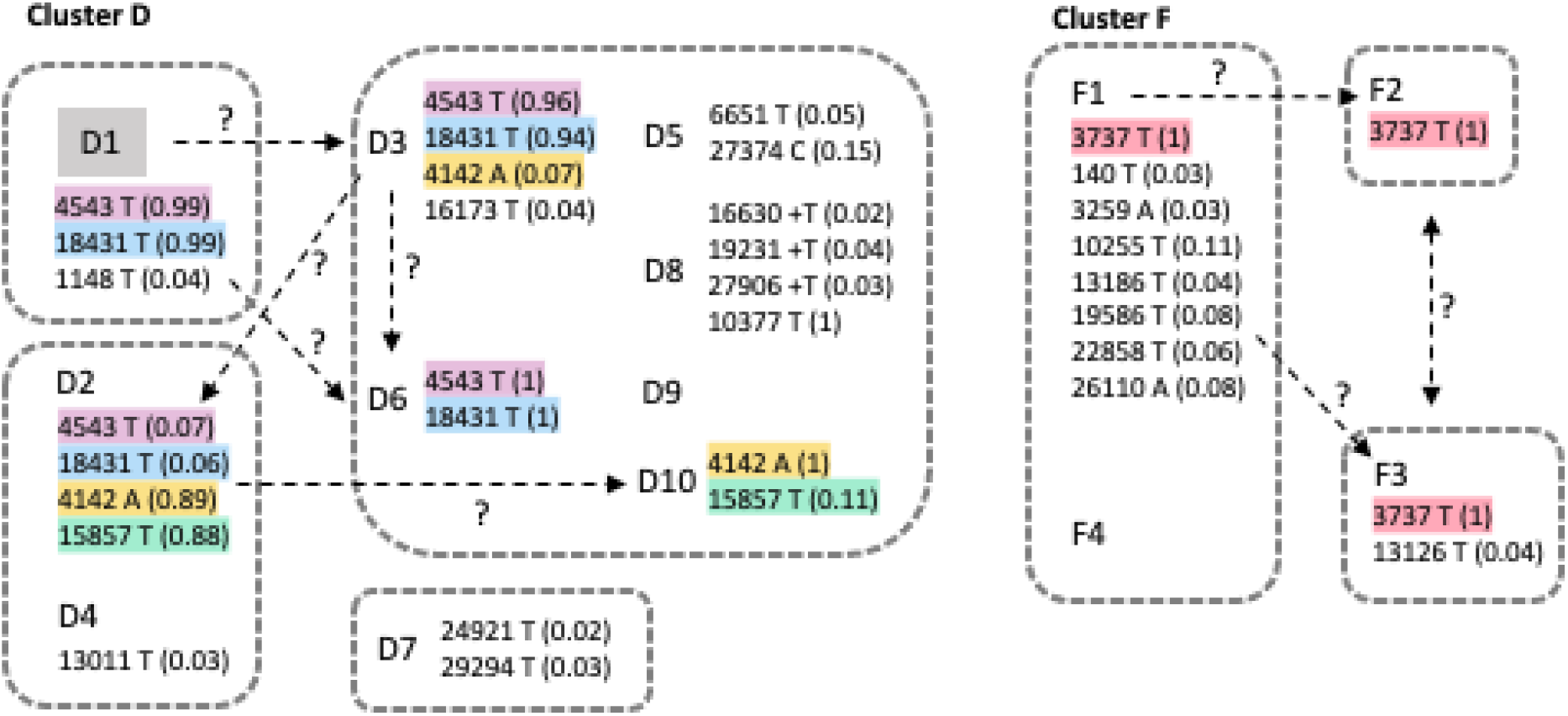
Shared variants and inferred transmission events for clusters D and F. Below each sample ID, we show variant site and allele, followed by its VAF in parenthesis. Fixed variants (VAF ≥ 0.98) shared by all members of a cluster are not shown. Dashed arrows indicate putative transmission events. Question marks highlight potential alternatives.

##### Statistical and graphical approaches

The Worby et al. approaches were not as helpful in inferring transmission as for delimiting clusters. Assigning the source of each sample to the patient with the highest number of shared intrahost variants or the minimum genetic distance (using weights or absolute values) resulted in samples with multiple potential sources and pairs with bidirectional transmission (**Figure 4A, B**). Relying only on intrahost shared variants proved inefficient, as most samples were not connected to any other (**Figure 4C**). Consensus sequences from the same cluster were very similar, so multiple samples were often equidistant, preventing choosing one of them as the source. The MST analysis (**Figure S4)** was incompatible with the epidemiological information. Apart from tied transmission paths for some of the clusters (clusters D and E, with three and two options, respectively), the starting point of the transmission did not coincide with the epidemiological information in any of the cases. TransPhylo could differentiate the different clusters (**Figure 7, Figure S5**); however, the inferred direct transmission events within clusters were often not compatible with the epidemiological information.

**Figure 7.**
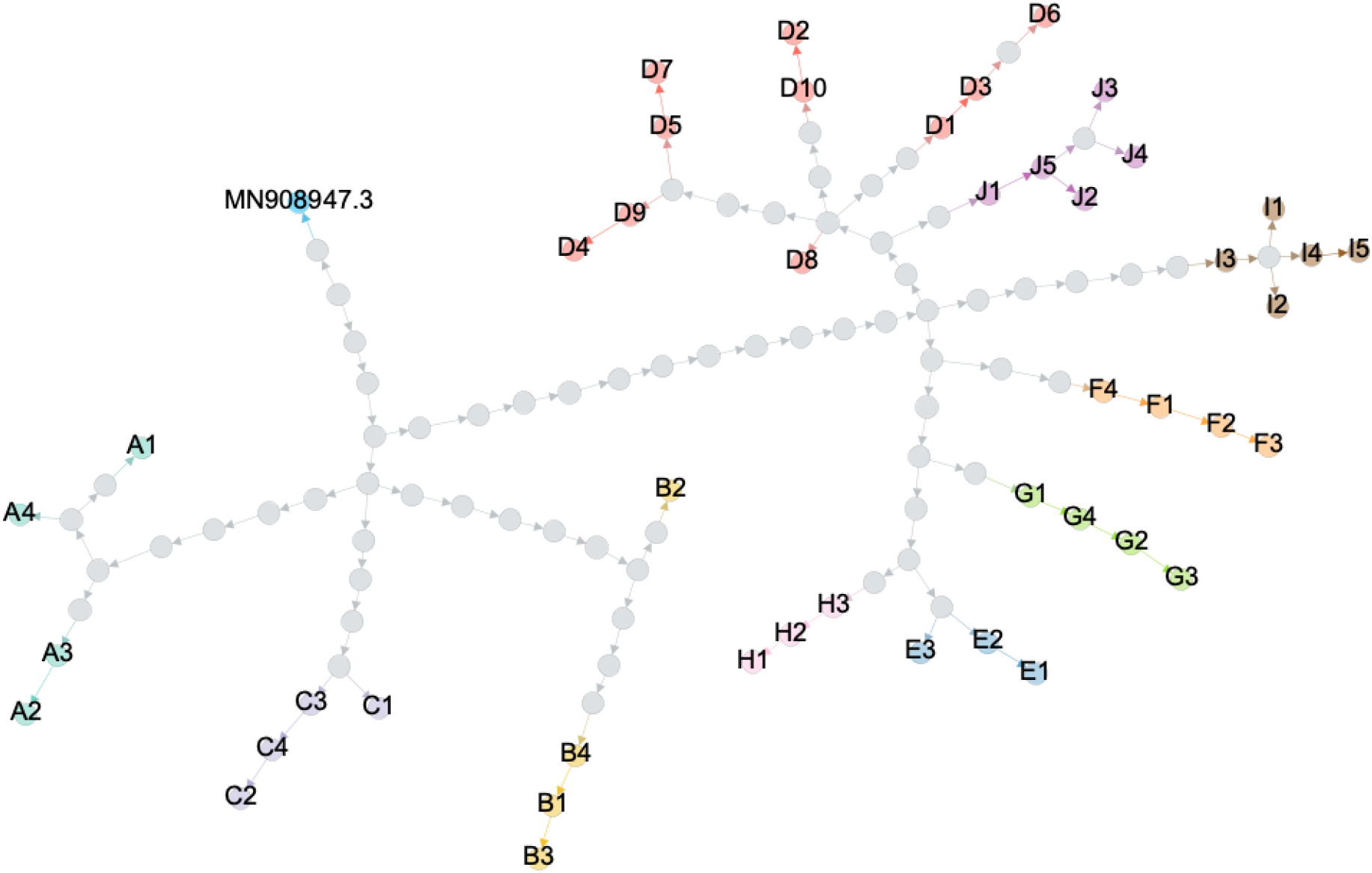
TransPhylo transmission graph. Gephi (Bastian et al. 2009) depiction of TransPhylo’s consensus transmission tree. Gray dots represent inferred unsampled individuals.

### Transmission bottleneck size

We selected individual pairs representing direct transmission events to estimate the transmission bottleneck size according to the epidemiological and genomic information. We discarded clusters A and B (nursing homes) from this analysis because it was impossible to identify likely transmission pairs in these cases. We had contact information about at least a transmission pair for clusters C, E, G, H, I, and J. The situation was more complex for clusters D and F, so we identified possible transmission pairs considering both the epidemiological and the genomic data, as described above.

Across the studied transmission pairs, we found an average of 0.38 (range 0-3) shared intrahost variants (**Table S6**). Accordingly, the estimated transmission bottleneck sizes were typically small (1-2 viral particles), except for the pair F1 → F3 (6 viral particles) (**Figure 8, Table S6**). To ensure that our selection of transmission pairs in clusters D and F was not biasing these estimates downwards, we also calculated the transmission bottleneck sizes for all potential pairs within these two clusters. The estimated bottlenecks were consistently 1-2. Note that the bottleneck size can only be estimated when there is at least one variant in the donor (regardless of whether that variant is observed in the recipient). If none of the donor variants appear in the recipient, the estimated bottleneck size will be 1, with a variable confidence interval depending on the variant calling threshold.

**Figure 8.**
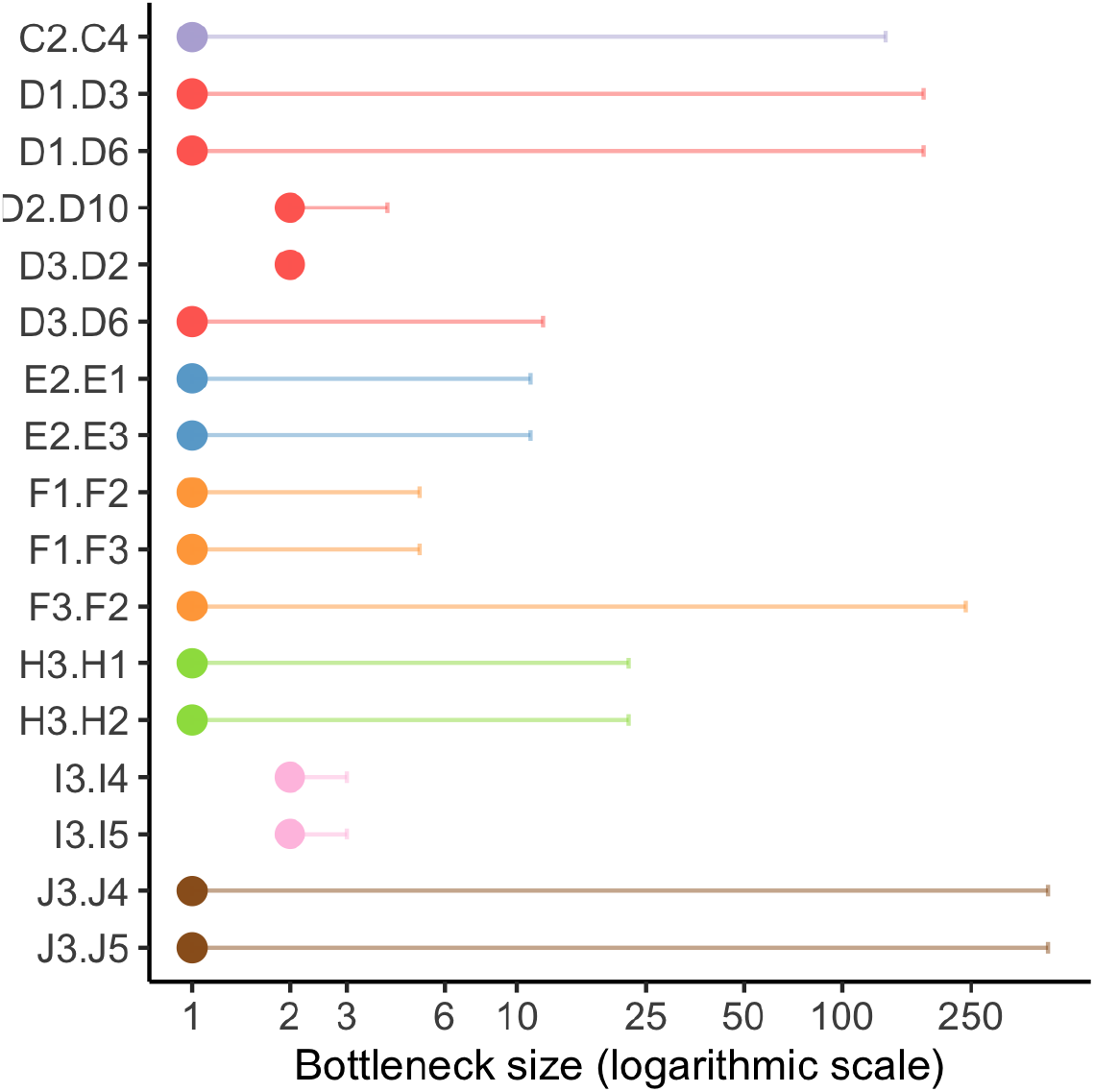
Estimated transmission bottleneck sizes. Labels on the Y-axis represent donor-recipient pairs. Estimates were obtained with the beta-binomial ML method (Sobel Leonard et al. 2017). Horizontal lines represent 95% confidence intervals. The X-axis is on a logarithmic scale.

**Figure 9.**
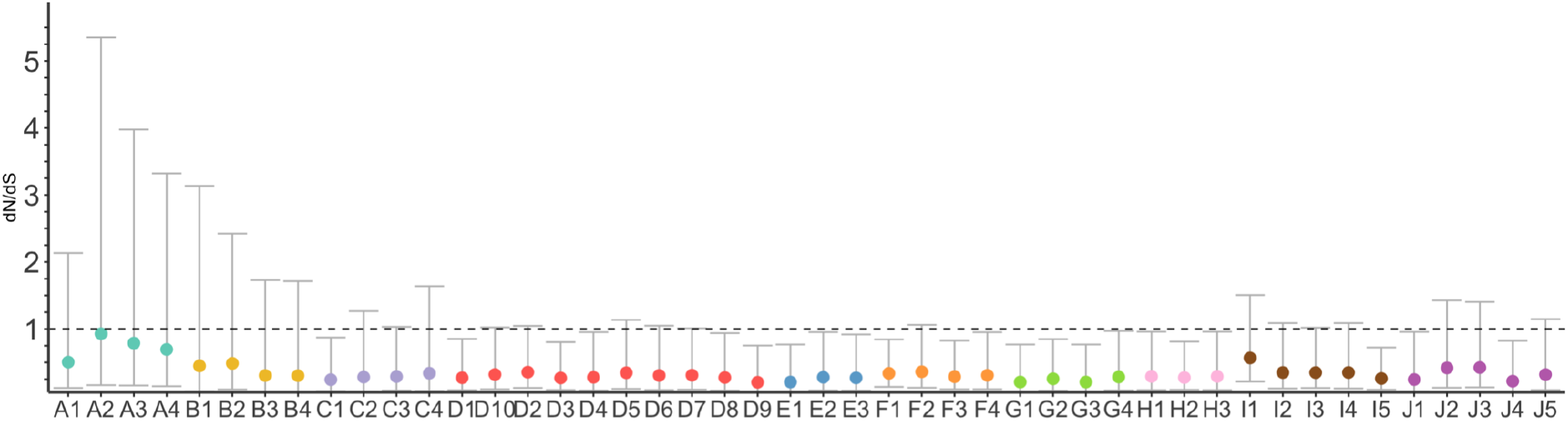
Estimated *dN/dS* values per sample for missense variants. Values were estimated using the dNdScv package. Vertical lines represent 95% confidence intervals.

### Assessment of selective pressures

We estimated *dN/dS* values for missense variants consistently below 1, suggesting a predominance of intrahost purifying selection across samples (**Figure 8**).

## Discussion

Understanding SARS-CoV-2 transmission is crucial to identify which situations minimize or maximize the risk of infection and, therefore, implement more effective control strategies. Previous studies have tried to reconstruct SARS-CoV-2 local transmission chains with more or less success using a combination of epidemiological and genomic data (Popa et al. 2020; Sekizuka et al. 2020; Shen et al. 2020; Hamilton et al. 2021; San et al. 2021). However, it is unclear whether, in situations for which contact tracing information is limited, we can use SARS-CoV-2 genomic information alone to understand who infected whom. Here, we show that while SARS-CoV-2 genomic variation can be helpful to delimit distinct transmission clusters, it might not be enough to resolve with confidence direct transmission events. Using the most likely transmission pairs, we infer a tiny transmission bottleneck size for SARS-CoV-2 in the order of 1-10 viral particles.

The level of interhost genomic variation that we detected was generally low. However, this did not prevent the distinction among local clusters sampled in the same month in the same city. When the sampling dates were taken into account, the concordance between genomic and epidemiological clusters was maximized, highlighting the relevance of the temporal information. Methods for cluster delimitation that rely exclusively on intrahost variants did not work well in this regard. In contrast, methods based on differences at the consensus level could differentiate the clusters near perfectly. The latter suggests that in SARS-CoV-2, consensus sequences alone are enough to separate samples belonging to different clusters from the same area. At the same time, intra-host diversity does not seem to be sufficient for this task.

We found a limited number of intrahost variants (∼8 before filtering recurrent variants and ∼3 after filtering), as reported in other studies (Kuipers et al. 2020; Seemann et al. 2020; Shen et al. 2020; Tonkin-Hill et al. 2020; Wölfel et al. 2020; Butler et al. 2021; Valesano et al. 2021; Y. Wang et al. 2021). Half of our samples (27/48) had a viral load above 10^3^ copies / µL, which is the threshold determined in Valesano et al. (2021) for reliable identification of intrahost variants with a VAF ≥ 2% in single replicates. Unlike previous studies, we used technical replicates to stress variant calling reproducibility and added unique barcodes to each sample to discard the potential effect of cross-sample contamination.

Here, transmission history within nursing homes or households, where most SARS-CoV-2 infections occur (Lee et al. 2020), was complicated to decipher. In general, all the methods we tried, even those relying on intrahost variation, could not provide clear transmission patterns within clusters, as seen before in care homes (Hamilton et al. 2021). The latter can be explained by lack of genetic variation but also by homoplasy, as we observed several shared intrahost variants among apparently unrelated samples. In addition, we noticed that if VAF thresholds are relaxed, unique or uncommon mutations appear in many individuals, suggesting these variants are either recurrent artifacts or correspond to hotspot mutations (Tonkin-Hill et al. 2020). Deciding which shared intrahost variants in SARS-CoV-2 are the result of transmission events is not easy. Much care should be taken regarding reliable genotyping and identifying recurrent events, particularly for samples with low viral loads (van Dorp, Acman, et al. 2020; van Dorp, Richard, et al. 2020; Kubik et al. 2021; Valesano et al. 2021). Recurrent, low-frequency insertions in SARS-CoV-2 have already been detected elsewhere (Kuipers et al. 2020; Rayko and Komissarov 2020; Tonkin-Hill et al. 2020; Turakhia et al. 2020).

Although not addressed in this study, another potential complication regarding the identification of shared intrahost variants is the occurrence of significant intrahost evolution. Several studies have reported VAF changes within days in SARS-CoV-2 (Jary et al. 2020; Tonkin-Hill et al. 2020; Voloch et al. 2020; Y. Wang et al. 2021), even faster in immunocompromised individuals (Avanzato et al. 2020; Kemp et al. 2021). On the other hand, more rigorous studies report that diversity does not increase over time, although this does not imply that VAFs cannot change significantly among different time points (Valesano et al. 2021). Significant intrahost evolution would imply that the amount of sharing between samples could change depending on the exact sampling dates, so the inferences derived from it.

Our estimates indicate that the SARS-CoV-2 transmission bottleneck size is small or very small, with only a few viral particles being responsible for the successful growth within the recipient, which is consistent with previous studies (Li et al. 2021; Lythgoe et al. 2021; San et al. 2021; D. Wang et al. 2021). Notably, minimal bottleneck estimates have also been obtained for a highly transmissible SARS-CoV-2 lineage like Delta (Li et al. 2021). In contrast, Popa et al. (2020) estimated an average transmission bottleneck size for SARS-CoV-2 of 1,000, but these estimates have been put into question (Martin and Koelle 2021). The advantages of our study in this regard are the technical replicates, cross-contamination controls, and consistent filters for recurrent variants. A small transmission bottleneck size for SARS-CoV-2 is consistent with a dominance of aerosol transmission over direct contact, as seen in influenza (Varble et al. 2014; Frise et al. 2016; McCrone and Lauring 2018).

If only one or a few virions are passed during transmission, then most of SARS-CoV-2 intrahost variation has to be due to the accumulation of *de novo* mutations (Voloch et al. 2020; Valesano et al. 2021). We inferred strong intrahost purifying selection across the genome for missense variants, as in previous comprehensive studies of SARS-CoV-2 intrahost variation (Tonkin-Hill et al. 2020). The latter is consistent with a severe transmission bottleneck reducing the efficiency of positive selection within hosts (McCrone and Lauring 2018).

In summary, our results suggest that SARS-CoV-2 genomic diversity is helpful to delimitate different transmission clusters within a relatively small area, but that could be insufficient to fully resolve transmissions within a household or in the same social event. Thus, contact tracing data will be essential to study direct SARS-CoV-2 transmission events, as it occurs in typical slow-evolving pathogens (Campbell et al. 2018; Campbell et al. 2019).

## Data Availability

Raw FASTQ files will be deposited at the National Center for Biotechnology Information (NCBI) Sequence Read Archive (SRA) (Leinonen et al. 2011). Viral consensus genomes will be available at the Global Initiative on Sharing All Influenza Data (GISAID) (Shu and McCauley 2017). Contact authors for details.

## Acknowledgments

This project was funded by grant EPICOVIGAL FONDO SUPERA-COVID19 from Banco Santander-CSIC-CRUE, grant CT850A-2 from ACIS SERGAS from the Consellería de Sanidade Xunta de Galicia, and grant ED431C2018/54-GRC from the Consellería de Cultura, Educación e Ordenación Universitaria of Xunta de Galicia. NS and TT were supported in part by a C3.ai Digital Transformation Institute award.

## Data availability

Raw FASTQ files have been deposited at the National Center for Biotechnology Information (NCBI) Sequence Read Archive (SRA) (Leinonen et al. 2011) (Project Accession No. PRJNAXXXXXX). Viral consensus genomes are available at the Global Initiative on Sharing All influenza Data (GISAID) (Shu and McCauley 2017) (accessions in Supplementary Table Sx).

## Author contributions

DP conceived the study and designed the analyses. SP, JJC, VdC, and BR obtained the patient samples and the epidemiological information. SP, NEG, LdC, IFS, and DV planned and performed the laboratory experiments. PGG, NV, NS, and TT carried out the bioinformatic analyses. DP wrote the draft manuscript with the help of PGG and NV. All authors read the manuscript and contributed to interpretation and discussion.

## Supplementary Data

Supplementary data are available at Virus Evolution online.

## Supplementary Material

**Table S1.** Transmission clusters. Cluster characteristics, sample ID, date of sampling, qPCR cycle threshold (Ct) (gene E) in the nasopharyngeal sample, viral load (copies / µL) (gene E) in the 1/10 diluted RNA extract, mean coverage and standard deviation (SD), and NextClade clade and PANGO lineage for each sample. Asterisks highlight discarded samples with poor sequencing quality.

| Cluster | Type | Sample ID | Ct | Viral load | Mean coverage | SD coverage | clade | lineage |
| --- | --- | --- | --- | --- | --- | --- | --- | --- |
| A | Nursing home | A1 | 17.6 | 2.18E+05 | 5299 | 2333 | 20A | B.1 |
|  |  | A2 | 17.3 | 2.32E+05 | 5690 | 255 | 20A | B.1 |
|  |  | A3 | 16.6 | 6.95E+05 | 5967 | 2557 | 20A | B.1 |
|  |  | A4 | 18.8 | 1.52E+05 | 6520 | 3034 | 20A | B.1 |
| B | Nursing home | B1 | 22.4 | 9.41E+03 | 5782 | 2981 | 20A | B.1 |
|  |  | B2 | 27.7 | 1.10E+02 | 4627 | 3029 | 20A | B.1 |
|  |  | B3 | 24.5 | 1.04E+03 | 5213 | 3046 | 20A | B.1 |
|  |  | B4 | 19.3 | 2.16E+05 | 6378 | 2836 | 20A | B.1 |
| C | Family | C1 | 21.8 | 4.17E+03 | 5550 | 3009 | 20A | B.1 |
|  |  | C2 | 17.3 | 2.71E+04 | 5918 | 2944 | 20A | B.1 |
|  |  | C3 | 19.4 | 2.90E+04 | 6955 | 3307 | 20A | B.1 |
|  |  | C4 | 31.6 | 7.85E+02 | 461 | 360 | 20A | B.1 |
| D | Birthday party | D1 | 16.6 | 6.32E+05 | 6290 | 2605 | 20E (EU1) | B.1.177 |
|  |  | D2 | 24.3 | 2.59E+03 | 6077 | 3041 | 20E (EU1) | B.1.177 |
|  |  | D3 | 29.7 | 3.57E+01 | 3852 | 2564 | 20E (EU1) | B.1.177 |
|  |  | D4 | 25.6 | 3.39E+02 | 6134 | 3474 | 20E (EU1) | B.1.177 |
|  |  | D5 | 19.0 | 4.55E+04 | 5958 | 2576 | 20E (EU1) | B.1.177 |
|  |  | D6 | 32.0 | 5.41E+00 | 953 | 844 | 20E (EU1) | B.1.177 |
|  |  | D7 | 23.0 | 1.21E+03 | 5407 | 2814 | 20E (EU1) | B.1.177 |
|  |  | D8 | 29.6 | 6.60E+00 | 1713 | 1381 | 20E (EU1) | B.1.177 |
|  |  | D9 | 19.7 | 1.30E+04 | 6192 | 2840 | 20E (EU1) | B.1.177 |
|  |  | D10 | 17.5 | 3.25E+04 | 5696 | 2397 | 20E (EU1) | B.1.177 |
|  |  | *D11 | 34.8 | 4.67E+00 | - | - | - | - |
| E | Family | E1 | 20.0 | 3.42E+04 | 6838 | 2917 | 20E (EU1) | B.1.177 |
|  |  | E2 | 25.8 | 1.79E+02 | 5112 | 2910 | 20E (EU1) | B.1.177 |
|  |  | E3 | 18.7 | 9.09E+04 | 6022 | 2519 | 20E (EU1) | B.1.177 |
| F | Family | F1 | 17.9 | 8.05E+04 | 5405 | 2171 | 20E (EU1) | B.1.177 |
|  |  | F2 | 24.3 | 1.25E+03 | 6172 | 3216 | 20E (EU1) | B.1.177 |
|  |  | F3 | 15.9 | 2.16E+05 | 6230 | 2553 | 20E (EU1) | B.1.177 |
|  |  | F4 | 21.0 | 1.26E+04 | 6781 | 3037 | 20E (EU1) | B.1.177 |
| G | Family | G1 | 16.6 | 4.39E+05 | 10796 | 3195 | 20E (EU1) | B.1.177 |
|  |  | G2 | 23.8 | 7.13E+02 | 10201 | 3820 | 20E (EU1) | B.1.177 |
|  |  | G3 | 28.0 | 3.99E+01 | 8252 | 4635 | 20E (EU1) | B.1.177 |
|  |  | G4 | 21.4 | 6.66E+03 | 10176 | 3827 | 20E (EU1) | B.1.177 |
|  |  | *G5 | 34.2 | 4.43E+00 | - | - | - | - |
| H | Family | H1 | 26.4 | 3.19E+01 | 5307 | 3515 | 20E (EU1) | B.1.177 |
|  |  | H2 | 23.6 | 2.28E+02 | 8241 | 4262 | 20E (EU1) | B.1.177 |
|  |  | H3 | 25.3 | 2.12E+01 | 1430 | 1140 | 20E (EU1) | B.1.177 |
| I | Birthday party/ family | I1 | 21.1 | 1.17E+03 | 4485 | 3120 | 20E (EU1) | B.1.177 |
|  |  | I2 | 26.4 | 1.93E+02 | 7989 | 4253 | 20E (EU1) | B.1.177 |
|  |  | I3 | 21.1 | 1.03E+05 | 9760 | 3514 | 20E (EU1) | B.1.177 |
|  |  | I4 | 22.2 | 3.33E+02 | 8378 | 4604 | 20E (EU1) | B.1.177 |
|  |  | I5 | 21.8 | 1.00E+03 | 8756 | 3931 | 20E (EU1) | B.1.177 |
|  |  | *I6 | 32 | 5.95E+00 | - | - | - | - |
| J | Family | J1 | 20.4 | 9.66E+02 | 9541 | 4502 | 20E (EU1) | B.1.177 |
|  |  | J2 | 24.0 | 8.94E+01 | 7573 | 4109 | 20E (EU1) | B.1.177 |
|  |  | J3 | 28.8 | 5.85E+01 | 5435 | 3146 | 20E (EU1) | B.1.177 |
|  |  | J4 | 21.8 | 8.63E+02 | 9465 | 4647 | 20E (EU1) | B.1.177 |
|  |  | J5 | 21.9 | 3.00E+03 | 9593 | 3968 | 20E (EU1) | B.1.177 |

**Table S2.** Sequencing quality control of each sample (both replicates). Red-colored samples that we excluded for bad quality. *Reads main barcode* is the number of reads mapped to the main barcode. *% main barcode* is the percentage of reads mapped to the main barcode. *Reads other barcodes* is the number of reads mapped to other barcodes. *% other barcode* is the percentage of reads mapped to another barcode. *Main barcode* is the identifier of the main barcode. *Mean coverage* is the mean coverage of reads mapped to reference. *Breadth 1/10/100X*: is the proportion of positions covered by at least 1/10/100 reads. Samples marked with an asterisk failed during sequencing.

| Sample name | Reads main barcode | % main barcode | Main barcode | Reads other barcodes | % other barcode | Mean coverage | Breadth 1X | Breadth 10X | Breadth 100X |
| --- | --- | --- | --- | --- | --- | --- | --- | --- | --- |
| A1-r1 | 2624 | 0.30 | A7 | 0 | 0 | 3152 | 1.000 | 0.999 | 0.997 |
| A1-r2 | 1226 | 0.20 | B3 | 0 | 0 | 2147 | 1.000 | 0.999 | 0.992 |
| A2-r1 | 1696 | 0.18 | B7 | 0 | 0 | 3286 | 1.000 | 0.999 | 0.998 |
| A2-r2 | 1298 | 0.19 | D3 | 0 | 0 | 2410 | 1.000 | 0.999 | 0.992 |
| A3-r1 | 656 | 0.08 | D7 | 0 | 0 | 2950 | 1.000 | 0.999 | 0.998 |
| A3-r2 | 734 | 0.09 | F3 | 2 | 0 | 3025 | 1.000 | 0.999 | 0.994 |
| A4-r1 | 2768 | 0.30 | E7 | 0 | 0 | 3181 | 1.000 | 0.999 | 0.993 |
| A4-r2 | 1492 | 0.15 | G3 | 2 | 0 | 3384 | 1.000 | 0.999 | 0.991 |
| B1-r1 | 3876 | 0.44 | F7 | 0 | 0 | 2929 | 1.000 | 0.999 | 0.992 |
| B1-r2 | 5458 | 0.64 | H3 | 0 | 0 | 2869 | 1.000 | 0.999 | 0.985 |
| B2-r1 | 18076 | 1.93 | H7 | 0 | 0 | 2649 | 1.000 | 0.999 | 0.992 |
| B2-r2 | 33834 | 3.64 | A4 | 8 | 0.001 | 2016 | 0.999 | 0.998 | 0.922 |
| B3-r1 | 9114 | 1.20 | A8 | 0 | 0 | 2477 | 1.000 | 0.999 | 0.983 |
| B3-r2 | 15370 | 1.66 | B4 | 2 | 0 | 2779 | 1.000 | 0.999 | 0.969 |
| B4-r1 | 2294 | 0.27 | B8 | 0 | 0 | 3095 | 1.000 | 0.999 | 0.998 |
| B4-r2 | 2274 | 0.24 | C4 | 2 | 0 | 3312 | 1.000 | 0.999 | 0.994 |
| C1-r1 | 5624 | 0.68 | G8 | 2 | 0 | 2695 | 1.000 | 0.999 | 0.991 |
| C1-r2 | 11252 | 1.18 | A5 | 0 | 0 | 2893 | 1.000 | 0.999 | 0.976 |
| C2-r1 | 3700 | 0.42 | E8 | 0 | 0 | 2999 | 1.000 | 0.999 | 0.992 |
| C2-r2 | 3010 | 0.35 | G4 | 0 | 0 | 2938 | 1.000 | 0.999 | 0.991 |
| C3-r1 | 3294 | 0.36 | F8 | 0 | 0 | 3225 | 1.000 | 0.999 | 0.993 |
| C3-r2 | 3840 | 0.35 | H4 | 2 | 0 | 3795 | 1.000 | 0.999 | 0.992 |
| C4-r1 | 68002 | 8.89 | C8 | 0 | 0 | 347 | 0.992 | 0.977 | 0.820 |
| C4-r2 | 66094 | 7.58 | F4 | 0 | 0 | 113 | 0.917 | 0.753 | 0.396 |
| D1-r1 | 1070 | 0.13 | E3 | 2 | 0 | 3000 | 1.000 | 0.999 | 0.999 |
| D1-r2 | 1032 | 0.11 | B5 | 0 | 0 | 3305 | 1.000 | 0.999 | 0.998 |
| D2-r1 | 6636 | 0.81 | E4 | 0 | 0 | 2760 | 0.999 | 0.999 | 0.992 |
| D2-r2 | 10224 | 0.99 | C5 | 0 | 0 | 3356 | 1.000 | 0.999 | 0.991 |
| D3-r1 | 27042 | 2.99 | E5 | 0 | 0 | 2189 | 0.999 | 0.999 | 0.985 |
| D3-r2 | 42476 | 4.03 | D5 | 0 | 0 | 1666 | 0.999 | 0.987 | 0.898 |
| D4-r1 | 16700 | 1.83 | A1 | 0 | 0 | 2853 | 1.000 | 0.999 | 0.978 |
| D4-r2 | 18806 | 1.69 | F5 | 0 | 0 | 3353 | 1.000 | 0.999 | 0.978 |
| D5-r1 | 4110 | 0.48 | B1 | 0 | 0 | 3037 | 1.000 | 0.999 | 0.999 |
| D5-r2 | 3216 | 0.38 | G5 | 0 | 0 | 2924 | 1.000 | 0.999 | 0.999 |
| D6-r1 | 76334 | 9.17 | C1 | 0 | 0 | 608 | 0.997 | 0.953 | 0.849 |
| D6-r2 | 57482 | 7.61 | H5 | 2 | 0 | 344 | 0.979 | 0.925 | 0.726 |
| D7-r1 | 9970 | 1.22 | E1 | 0 | 0 | 2705 | 1.000 | 0.999 | 0.998 |
| D7-r2 | 10636 | 1.27 | B6 | 0 | 0 | 2711 | 0.999 | 0.999 | 0.990 |
| D8-r1 | 45386 | 5.89 | F1 | 0 | 0 | 1176 | 0.999 | 0.991 | 0.888 |
| D8-r2 | 56576 | 9.20 | C6 | 0 | 0 | 536 | 0.990 | 0.954 | 0.802 |
| D9-r1 | 4678 | 0.48 | G1 | 0 | 0 | 3361 | 1.000 | 0.999 | 0.998 |
| D9-r2 | 6650 | 0.79 | D6 | 0 | 0 | 2845 | 1.000 | 0.999 | 0.998 |
| D10-r1 | 3356 | 0.39 | H1 | 0 | 0 | 3090 | 1.000 | 0.999 | 0.999 |
| D10-r2 | 4044 | 0.55 | E6 | 0 | 0 | 2608 | 1.000 | 0.999 | 0.999 |
| D11-r1* | 78992 | 8.53 | D1 | 0 | 0 | 24 | 0.379 | 0.311 | 0.085 |
| D11-r2* | 65230 | 9.83 | A6 | 0 | 0 | 18 | 0.443 | 0.335 | 0.030 |
| E1-r1 | 6360 | 0.62 | A2 | 0 | 0 | 3539 | 1.000 | 0.999 | 0.999 |
| E1-r2 | 5576 | 0.57 | F6 | 0 | 0 | 3325 | 1.000 | 0.999 | 0.999 |
| E2-r1 | 14598 | 1.62 | B2 | 0 | 0 | 2942 | 1.000 | 0.999 | 0.988 |
| E2-r2 | 11042 | 1.65 | G6 | 0 | 0 | 2195 | 1.000 | 0.999 | 0.965 |
| E3-r1 | 3278 | 0.34 | C2 | 0 | 0 | 3340 | 1.000 | 0.999 | 0.999 |
| E3-r2 | 2668 | 0.34 | H6 | 0 | 0 | 2689 | 1.000 | 0.999 | 0.999 |
| F1-r1 | 2182 | 0.24 | D2 | 0 | 0 | 3216 | 1.000 | 0.999 | 0.999 |
| F1-r2 | 2252 | 0.36 | C7 | 0 | 0 | 2191 | 1.000 | 0.999 | 0.998 |
| F2-r1 | 12540 | 1.22 | E2 | 0 | 0 | 3390 | 1.000 | 0.999 | 0.998 |
| F2-r2 | 7492 | 0.86 | G7 | 0 | 0 | 2822 | 0.999 | 0.999 | 0.984 |
| F3-r1 | 1906 | 0.23 | F2 | 0 | 0 | 2806 | 1.000 | 0.999 | 0.999 |
| F3-r2 | 2898 | 0.30 | D8 | 0 | 0 | 3439 | 1.000 | 0.999 | 0.999 |
| F4-r1 | 4088 | 0.40 | G2 | 0 | 0 | 3588 | 1.000 | 0.999 | 0.999 |
| F4-r2 | 7258 | 0.77 | H8 | 0 | 0 | 3219 | 1.000 | 0.999 | 0.998 |
| G1-r1 | 2536 | 0.16 | D2 | 0 | 0 | 5427 | 1.000 | 1.000 | 0.999 |
| G1-r2 | 2666 | 0.15 | G4 | 6 | 0 | 6111 | 1.000 | 0.999 | 0.999 |
| G2-r1 | 16432 | 0.88 | A1 | 2 | 0 | 6027 | 1.000 | 0.999 | 0.999 |
| G2-r2 | 6634 | 0.45 | D3 | 8 | 0.001 | 4932 | 1.000 | 0.999 | 0.999 |
| G3-r1 | 46832 | 2.72 | B1 | 0 | 0 | 4382 | 1.000 | 0.999 | 0.985 |
| G3-r2 | 47348 | 2.81 | E3 | 24 | 0.001 | 4404 | 1.000 | 0.999 | 0.976 |
| G4-r1 | 8824 | 0.54 | C1 | 0 | 0 | 5468 | 1.000 | 0.999 | 0.999 |
| G4-r2 | 9356 | 0.58 | F3 | 0 | 0 | 5292 | 1.000 | 0.999 | 0.999 |
| G5-r1* | 223300 | 12.95 | D1 | 0 | 0 | 8 | 0.063 | 0.037 | 0.032 |
| G5-r2* | 171046 | 11.19 | G3 | 4 | 0 | 22 | 0.953 | 0.409 | 0.071 |
| H1-r1 | 88504 | 5.95 | F1 | 0 | 0 | 2617 | 1.000 | 0.999 | 0.967 |
| H1-r2 | 113642 | 7.16 | A4 | 32 | 0.002 | 2764 | 1.000 | 0.999 | 0.931 |
| H2-r1 | 36290 | 2.29 | C3 | 0 | 0 | 4529 | 1.000 | 1.000 | 0.999 |
| H2-r2 | 36534 | 2.37 | F5 | 0 | 0 | 4123 | 1.000 | 1.000 | 0.991 |
| H3-r1 | 191952 | 13.08 | E1 | 0 | 0 | 827 | 1.000 | 0.990 | 0.888 |
| H3-r2 | 180454 | 11.07 | H3 | 4 | 0 | 763 | 0.999 | 0.954 | 0.814 |
| I1-r1 | 84412 | 5.55 | H2 | 0 | 0 | 2105 | 1.000 | 0.999 | 0.969 |
| I1-r2 | 72380 | 4.67 | C5 | 22 | 0.001 | 2479 | 1.000 | 1.000 | 0.970 |
| I2-r1 | 30912 | 2.18 | E2 | 0 | 0 | 3714 | 1.000 | 0.999 | 0.992 |
| I2-r2 | 28066 | 1.60 | H4 | 10 | 0 | 4599 | 1.000 | 0.999 | 0.995 |
| I3-r1 | 5982 | 0.38 | C2 | 0 | 0 | 5261 | 1.000 | 0.999 | 0.999 |
| I3-r2 | 6212 | 0.42 | F4 | 2 | 0 | 4928 | 1.000 | 0.999 | 0.999 |
| I4-r1 | 37058 | 2.45 | B2 | 0 | 0 | 4299 | 1.000 | 1.000 | 0.999 |
| I4-r2 | 28434 | 1.83 | E4 | 4 | 0 | 4628 | 1.000 | 0.999 | 0.999 |
| I5-r1 | 15330 | 0.92 | A2 | 0 | 0 | 5003 | 1.000 | 0.999 | 0.999 |
| I5-r2 | 11476 | 0.82 | D4 | 124 | 0.009 | 4195 | 1.000 | 1.000 | 0.999 |
| I6-r1* | 222382 | 13.50 | G2 | 34 | 0.002 | 98 | 0.513 | 0.454 | 0.369 |
| I6-r2* | 191802 | 11.42 | B5 | 0 | 0 | 111 | 0.927 | 0.541 | 0.317 |
| J1-r1 | 26238 | 1.56 | F2 | 0 | 0 | 5015 | 1.000 | 1.000 | 0.999 |
| J1-r2 | 28080 | 1.59 | A5 | 20 | 0.001 | 5432 | 1.000 | 1.000 | 0.999 |
| J2-r1 | 75046 | 4.60 | A3 | 0 | 0 | 3780 | 1.000 | 0.999 | 0.984 |
| J2-r2 | 48592 | 3.07 | D5 | 2 | 0 | 4035 | 1.000 | 1.000 | 0.981 |
| J3-r1 | 32736 | 1.97 | H1 | 14 | 0.001 | 3152 | 1.000 | 0.999 | 0.983 |
| J3-r2 | 30096 | 2.35 | C4 | 6 | 0 | 2374 | 1.000 | 0.999 | 0.975 |
| J4-r1 | 34560 | 1.96 | B3 | 2 | 0 | 5187 | 1.000 | 0.999 | 0.999 |
| J4-r2 | 37946 | 2.11 | E5 | 0 | 0 | 5239 | 1.000 | 1.000 | 0.984 |
| J5-r1 | 12098 | 0.73 | G1 | 0 | 0 | 5330 | 1.000 | 0.999 | 0.999 |
| J5-r2 | 14738 | 0.97 | B4 | 16 | 0.001 | 4858 | 1.000 | 0.999 | 0.999 |

**Table S3.** Average number of differences among consensus sequences within and between clusters.

|  | **A** | **B** | **C** | **D** | **E** | **F** | **G** | **H** | **I** | **J** |
| --- | --- | --- | --- | --- | --- | --- | --- | --- | --- | --- |
| **A** | 0.5 |  |  |  |  |  |  |  |  |  |
| **B** | 2.75 | 1 |  |  |  |  |  |  |  |  |
| **C** | 2.5 | 2.75 | 0.5 |  |  |  |  |  |  |  |
| **D** | 7.25 | 7.5 | 7.25 | 1.69 |  |  |  |  |  |  |
| **E** | 6.25 | 6.5 | 6.25 | 1 | 0 |  |  |  |  |  |
| **F** | 11 | 11.25 | 11 | 5.75 | 4.75 | 0.5 |  |  |  |  |
| **G** | 7.25 | 7.5 | 7.25 | 2 | 1 | 5.75 | 0 |  |  |  |
| **H** | 7.25 | 7.5 | 7.25 | 2 | 1 | 5.75 | 2 | 0 |  |  |
| **I** | 7.45 | 7.7 | 7.45 | 2.2 | 1.2 | 5.95 | 2.2 | 2.2 | 0.4 |  |
| **J** | 6.25 | 6.5 | 6.25 | 1 | 0 | 4.75 | 1 | 1 | 1.2 | 0 |

**Table S4.**
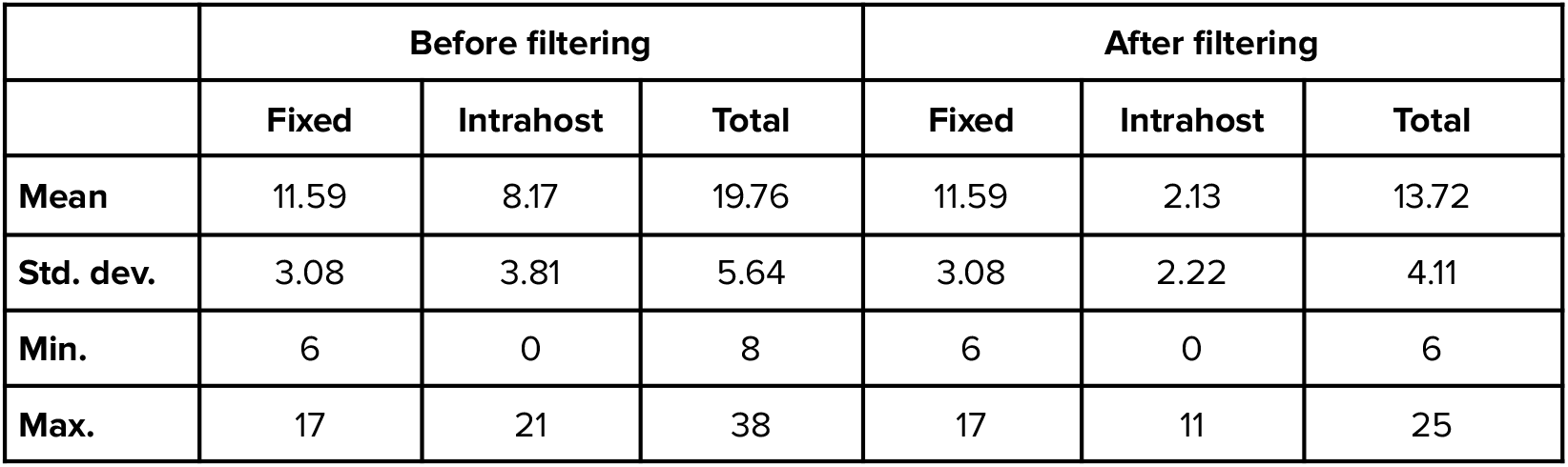
Number of intrahost and fixed variants (SNVs and indels) per sample.

|  | Before filtering |  |  | After filtering |  |  |
| --- | --- | --- | --- | --- | --- | --- |
|  | Fixed | Intrahost | Total | Fixed | Intrahost | Total |
| **Mean** | 11.59 | 8.17 | 19.76 | 11.59 | 2.13 | 13.72 |
| **Std. dev.** | 3.08 | 3.81 | 5.64 | 3.08 | 2.22 | 4.11 |
| **Min.** | 6 | 0 | 8 | 6 | 0 | 6 |
| **Max.** | 17 | 21 | 38 | 17 | 11 | 25 |

**Table S5.** Low-frequency variants (0.5 > VAF ≥ 0.02) evaluated. Number (#) and percentage (%) of samples in which each variant appears. Mean VAF and standard deviation (sd) of the variant and samples ID. The asterisk indicates variants with a VAF > 0.5 in at least another sample of the same cluster.

| Variant (VAF < 0.5) | # samples | % samples | mean VAF | sd | Samples that present the variant (VAF > 0.02) |
| --- | --- | --- | --- | --- | --- |
| **Low-frequency variants removed** |  |  |  |  |  |
| C6696+T | 39 | 84.78 | 0.064 | 0.014 | A1, A3, A4, B1, B2, B4, C1, C3, D1, D10, D2, D3, D4, D5, D7, D8, D9, E1, E2, E3, F1, F2, F3, F4, G1, G2, G3, G4, H1, H2, I2, I3, I4, I5, J1, J2, J3, J4, J5 |
| C29051+A | 39 | 84.78 | 0.038 | 0.008 | A1, A2, A3, A4, B1, B2, B3, B4, C1, C2, C3, D1, D10, D2, D3, D4, D5, D7, D9, E1, E2, E3, F1, F2, F3, F4, G1, G2, G3, G4, H2, I2, I3, I4, I5, J1, J2, J4, J5 |
| C19983+T | 30 | 65.22 | 0.033 | 0.006 | A1, B1, B2, B3, B4, C3, D1, D10, D2, D4, D5, D7, D9, E1, E2, E3, F1, F2, F3, F4, G1, G2, G3, G4, H2, I4, J1, J2, J4, J5 |
| G15965+T | 30 | 65.22 | 0.042 | 0.016 | A1, A2, A3, A4, B1, B2, B3, B4, C1, C2, C3, D1, D2, D3, D4, D7, E1, E2, E3, F1, F2, F3, F4, G1, G4, H2, I2, I3, I4, J5 |
| C28214+T | 26 | 56.52 | 0.026 | 0.004 | A2, A3, A4, B1, B3, B4, C1, C2, C3, D1, D10, D2, D4, D5, D9, E1, E3, F1, F2, F3, F4, G3, G4, I1, I3, I4 |
| C11074+T | 24 | 52.17 | 0.031 | 0.006 | A1, A3, A4, B1, B2, B3, B4, C1, C3, D1, D3, D7, D9, E2, E3, F2, G1, G3, G4, H1, I4, I5, J3, J5 |
| G21101+T | 14 | 30.43 | 0.033 | 0.016 | D10, D4, D5, D6, D8, D9, E2, F1, F2, F3, F4, G4, J2, J4 |
| C11812+A | 14 | 30.43 | 0.024 | 0.003 | A2, A3, B1, B3, B4, D1, D10, D2, D4, E2, F1, F2, F3, F4 |
| C10386+T | 12 | 26.09 | 0.023 | 0.003 | A3, D1, D10, D3, D4, D5, E2, E3, F1, F2, F3, F4 |
| G8927+T | 11 | 23.91 | 0.024 | 0.004 | B1, D10, D5, F1, F2, F3, F4, G3, H1, H2, J4 |
| G18368+T | 10 | 21.74 | 0.024 | 0.004 | A2, B1, B3, C1, C2, D1, D2, D3, F3, G3 |
| A29188G | 7 | 15.22 | 0.026 | 0.004 | D10, D5, D9, F1, F2, F3, F4 |
| C29187T | 7 | 15.22 | 0.026 | 0.004 | D10, D5, D9, F1, F2, F3, F4 |
| C27791+T | 6 | 13.04 | 0.028 | 0.005 | D10, D9, E2, F3, F4, J1 |
| C9502T | 6 | 13.04 | 0.022 | 0.001 | A3, B1, D4, E3, F1, F3 |
| G1730A | 6 | 13.04 | 0.023 | 0.002 | B1, B3, F1, F2, F3, J4 |
| C25703+T | 5 | 10.87 | 0.042 | 0.010 | A1, A2, A3, A4, J2 |
| C10619A | 4 | 8.70 | 0.021 | 0.000 | A3, D2, D3, F4 |
| C11095+T | 3 | 6.52 | 0.021 | 0.001 | B3, D9, F2 |
| C8905+T | 3 | 6.52 | 0.021 | 0.001 | D5, F1, F3 |
| C10619+A | 3 | 6.52 | 0.024 | 0.003 | B3, B4, C2 |
| G23903+T | 3 | 6.52 | 0.023 | 0.001 | F2, F4, G3 |
| G7889+T | 2 | 4.35 | 0.025 | 0.004 | D8, H1 |
| G26654+T | 2 | 4.35 | 0.021 | 0.001 | D3, F3 |
| C24358+A | 2 | 4.35 | 0.031 | 0.004 | B2, D8 |
| **Low-frequency variants kept** |  |  |  |  |  |
| T29834-C | 3 | 6.52 | 0.271 | 0.042 | H1, H2, H3 |
| C28748T | 1 | 2.17 | 0.040 |  | C3 |
| C28744T | 1 | 2.17 | 0.044 |  | H2 |
| T28867C | 1 | 2.17 | 0.135 |  | A1 |
| G13571T | 1 | 2.17 | 0.027 |  | J2 |
| G16246T | 1 | 2.17 | 0.036 |  | I5 |
| C6651T | 1 | 2.17 | 0.052 |  | D5 |
| C9570T | 1 | 2.17 | 0.021 |  | I1 |
| G13126T | 1 | 2.17 | 0.038 |  | F3 |
| G1148T | 1 | 2.17 | 0.042 |  | D1 |
| G10255T | 1 | 2.17 | 0.109 |  | F1 |
| C9264T | 1 | 2.17 | 0.020 |  | J3 |
| C7979T | 1 | 2.17 | 0.132 |  | B3 |
| G25218+T | 1 | 2.17 | 0.022 |  | H1 |
| G19231+T | 1 | 2.17 | 0.039 |  | D8 |
| G3639A | 1 | 2.17 | 0.024 |  | A4 |
| T28345A | 1 | 2.17 | 0.424 |  | A3 |
| T27374C | 1 | 2.17 | 0.150 |  | D5 |
| T1813+A | 1 | 2.17 | 0.022 |  | H1 |
| T1041C | 1 | 2.17 | 0.030 |  | I1 |
| G7312+T | 1 | 2.17 | 0.021 |  | G3 |
| G4232T | 1 | 2.17 | 0.092 |  | C1 |
| G4142A * | 1 | 2.17 | 0.066 |  | D3 |
| G3259A | 1 | 2.17 | 0.031 |  | F1 |
| G20126A | 1 | 2.17 | 0.039 |  | J4 |
| G29294T | 1 | 2.17 | 0.025 |  | D7 |
| G28001A | 1 | 2.17 | 0.061 |  | C1 |
| G27906+T | 1 | 2.17 | 0.034 |  | D8 |
| G25996T | 1 | 2.17 | 0.065 |  | G2 |
| G25555T | 1 | 2.17 | 0.270 |  | C3 |
| G25401A | 1 | 2.17 | 0.020 |  | H2 |
| C4543T * | 1 | 2.17 | 0.070 |  | D2 |
| G22104A | 1 | 2.17 | 0.022 |  | J3 |
| C5167T | 1 | 2.17 | 0.029 |  | I5 |
| A13154-TG | 1 | 2.17 | 0.038 |  | I1 |
| C3653T | 1 | 2.17 | 0.372 |  | J2 |
| C18175T | 1 | 2.17 | 0.029 |  | E3 |
| C16650T | 1 | 2.17 | 0.032 |  | B2 |
| C16630+T | 1 | 2.17 | 0.022 |  | D8 |
| C16173T | 1 | 2.17 | 0.040 |  | D3 |
| C15857T* | 1 | 2.17 | 0.105 |  | D10 |
| C14423T | 1 | 2.17 | 0.023 |  | I1 |
| C140T | 1 | 2.17 | 0.026 |  | F1 |
| C13667+T | 1 | 2.17 | 0.031 |  | H1 |
| C13541+T | 1 | 2.17 | 0.033 |  | H1 |
| C13011T | 1 | 2.17 | 0.028 |  | D4 |
| C11913T | 1 | 2.17 | 0.027 |  | I1 |
| C11074-T | 1 | 2.17 | 0.020 |  | I3 |
| C10790T | 1 | 2.17 | 0.022 |  | J2 |
| C10765T | 1 | 2.17 | 0.047 |  | C2 |
| A9026+T | 1 | 2.17 | 0.035 |  | H1 |
| A5146G | 1 | 2.17 | 0.108 |  | C3 |
| A28612G | 1 | 2.17 | 0.051 |  | J4 |
| A21625T | 1 | 2.17 | 0.067 |  | I3 |
| A18259G | 1 | 2.17 | 0.059 |  | G2 |
| A14185-G | 1 | 2.17 | 0.028 |  | I1 |
| C1701+T | 1 | 2.17 | 0.030 |  | H1 |
| C18431T* | 1 | 2.17 | 0.063 |  | D2 |
| C3330T | 1 | 2.17 | 0.042 |  | C1 |
| C18441T | 1 | 2.17 | 0.063 |  | I5 |
| C29686T | 1 | 2.17 | 0.033 |  | B3 |
| A13186T | 1 | 2.17 | 0.041 |  | F1 |
| C28005T | 1 | 2.17 | 0.455 |  | A3 |
| C27925T | 1 | 2.17 | 0.220 |  | H2 |
| C27532A | 1 | 2.17 | 0.206 |  | E2 |
| C26110A | 1 | 2.17 | 0.079 |  | F1 |
| C25797+A | 1 | 2.17 | 0.021 |  | F3 |
| C25427T | 1 | 2.17 | 0.338 |  | C3 |
| C24921T | 1 | 2.17 | 0.023 |  | D7 |
| C23816-T | 1 | 2.17 | 0.021 |  | I1 |
| C23202T | 1 | 2.17 | 0.112 |  | G4 |
| C22858T | 1 | 2.17 | 0.057 |  | F1 |
| C22713T | 1 | 2.17 | 0.033 |  | J5 |
| C21732T | 1 | 2.17 | 0.020 |  | J3 |
| C21575T | 1 | 2.17 | 0.031 |  | J5 |
| C2146T | 1 | 2.17 | 0.446 |  | B4 |
| C19586T | 1 | 2.17 | 0.083 |  | F1 |
| C19290T | 1 | 2.17 | 0.026 |  | I5 |
| C19269T | 1 | 2.17 | 0.041 |  | C1 |
| T5254G | 1 | 2.17 | 0.020 |  | I1 |

**Table S6.** Estimated transmission bottleneck sizes. The table shows the number and type of variants used for the calculation of the transmission bottleneck size. CI indicates the 95% likelihood confidence interval. Shared intrahost variants can be fixed in the recipient but not in the donor. Private intrahost donor variants are those that appear in the donor but not in the recipient. “-”: indicates pairs in which the absence of intrahost donor variants precludes the calculation of the bottleneck size.

| Cluster | Transmission pair | Bottleneck size | 95% CI | Private intrahost donor variants | Shared intrahost variants |
| --- | --- | --- | --- | --- | --- |
| C | C2.C4 | 1 | 1-136 | 1 | 0 |
| D | D1.D3 | 1 | 1-178 | 1 | 0 |
|  | D1.D6 | 1 | 1-178 | 1 | 0 |
|  | D2.D10 | 2 | 2-4 | 3 | 1 |
|  | D3.D2 | 2 | 2-2 | 1 | 3 |
|  | D3.D6 | 1 | 1-12 | 4 | 0 |
| E | E1.E2 | NA | NA | 0 | 0 |
|  | E1.E3 | NA | NA | 0 | 0 |
|  | E2.E1 | 1 | 1-11 | 1 | 0 |
|  | E2.E3 | 1 | 1-11 | 1 | 0 |
| F | F1.F2 | 1 | 1-5 | 7 | 0 |
|  | F1.F3 | 1 | 1-5 | 7 | 0 |
|  | F2.F3 | NA | NA | 0 | 0 |
|  | F3.F2 | 1 | 1-240 | 1 | 0 |
| G | G1.G2 | NA | NA | 0 | 0 |
|  | G1.G3 | NA | NA | 0 | 0 |
|  | H3.H1 | 1 | 1-22 | 4 | 0 |
|  | H3.H2 | 1 | 1-22 | 4 | 0 |
|  | I3.I4 | 2 | 2-3 | 1 | 3 |
|  | I3.I5 | 2 | 2-3 | 1 | 3 |
| J | J1.J2 | NA | NA | 0 | 0 |
|  | J1.J3 | NA | NA | 0 | 0 |
|  | J1.J4 | NA | NA | 0 | 0 |
|  | J1.J5 | NA | NA | 0 | 0 |
|  | J3.J4 | 1 | 1-430 | 3 | 0 |
|  | J3.J5 | 1 | 1-430 | 3 | 0 |
| average |  | 1.24 | 2.34-162.62 | 1.69 | 0.38 |
| sd |  | 0.44 | 3.67-229.77 | 2.07 | 0.98 |
| median |  | 1 | 1-12 | 1 | 0 |

**Figure S1.**
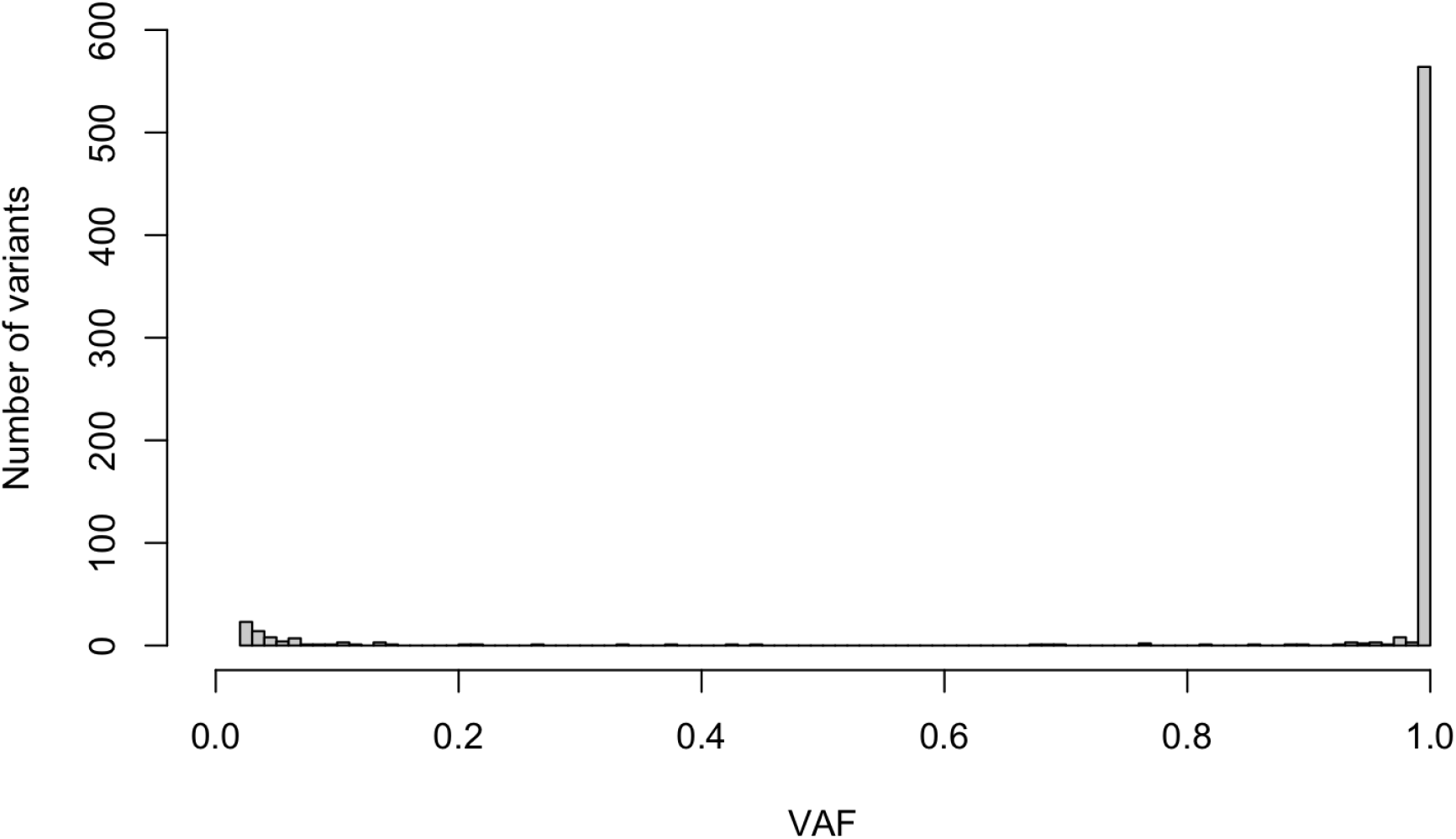
Variant allele frequency (VAF) distribution across samples. VAFs were calculated after filtering recurrent low-frequency variants.

**Figure S2.**
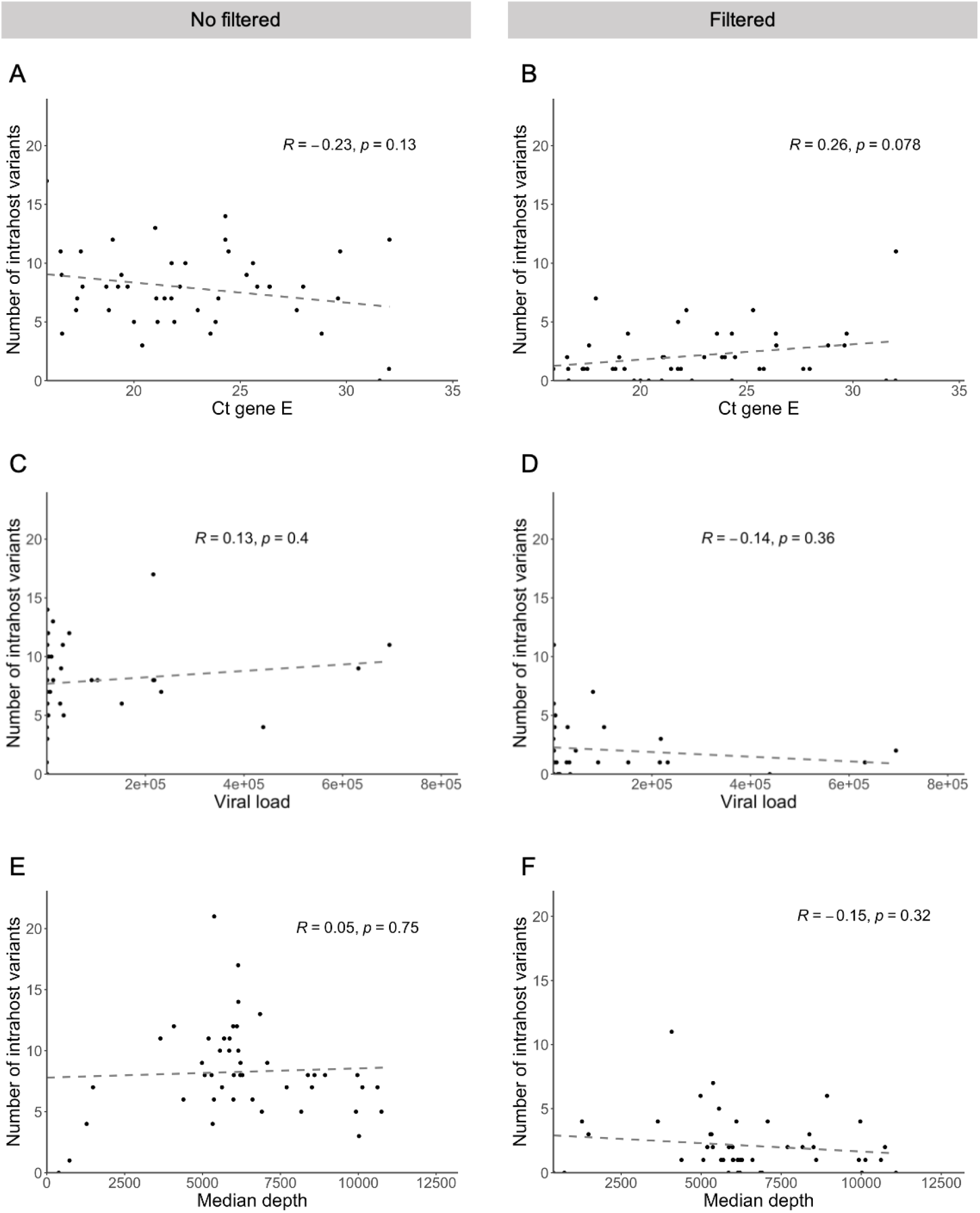
Number of intrahost variants. The plots describe the variation of the number of intrahost variants regarding Ct values (A, B), viral load (C, D), and sequencing depth (E, F), before (A, C, E) and after filtering recurrent low-frequency variants (B, D, F).

**Figure S3.**
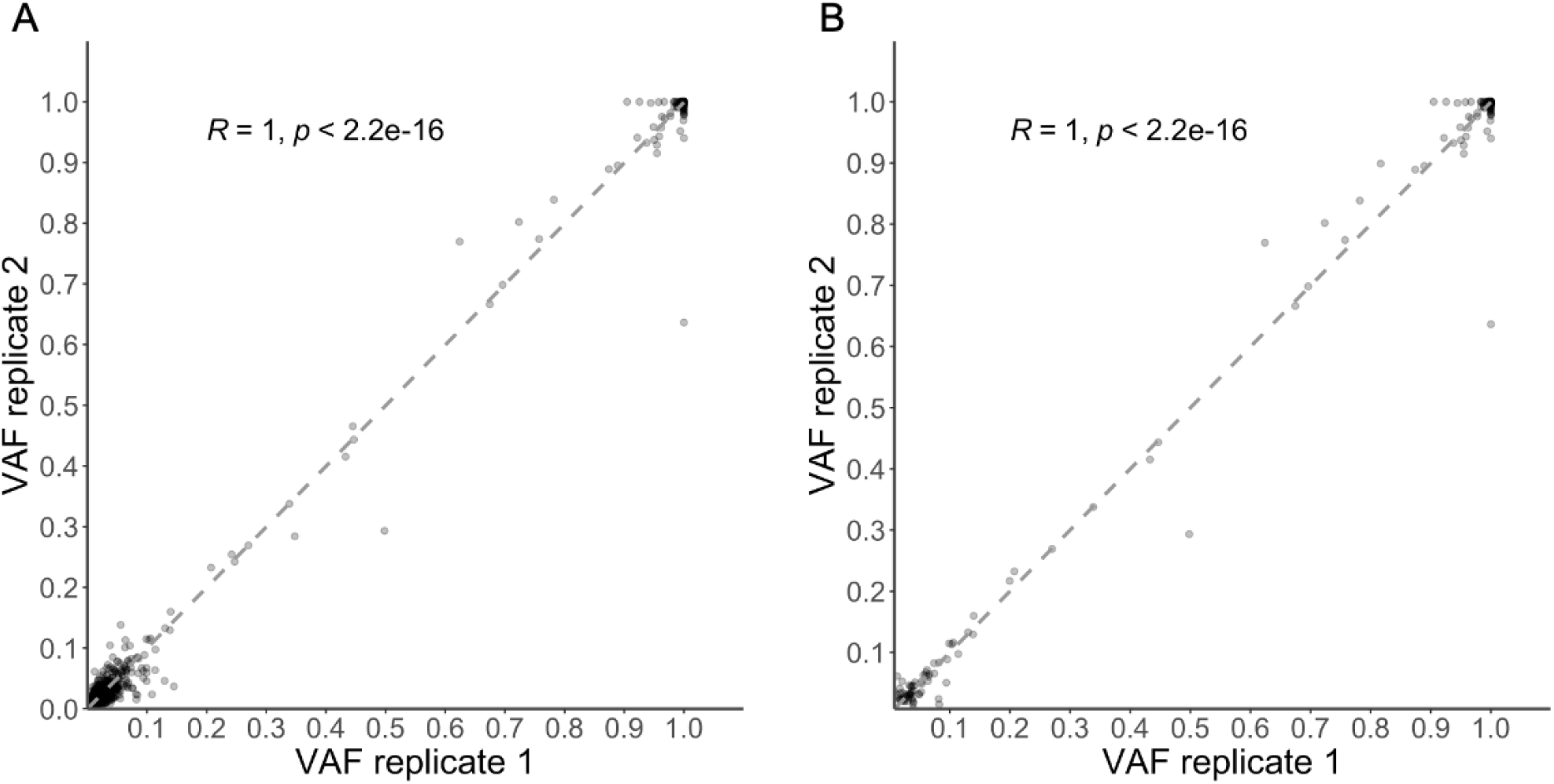
VAF correlation (Pearson) between replicates across samples. (A) after masking and before filtering recurrent low-frequency variants. (B) after masking and filtering.

**Figure S4.**
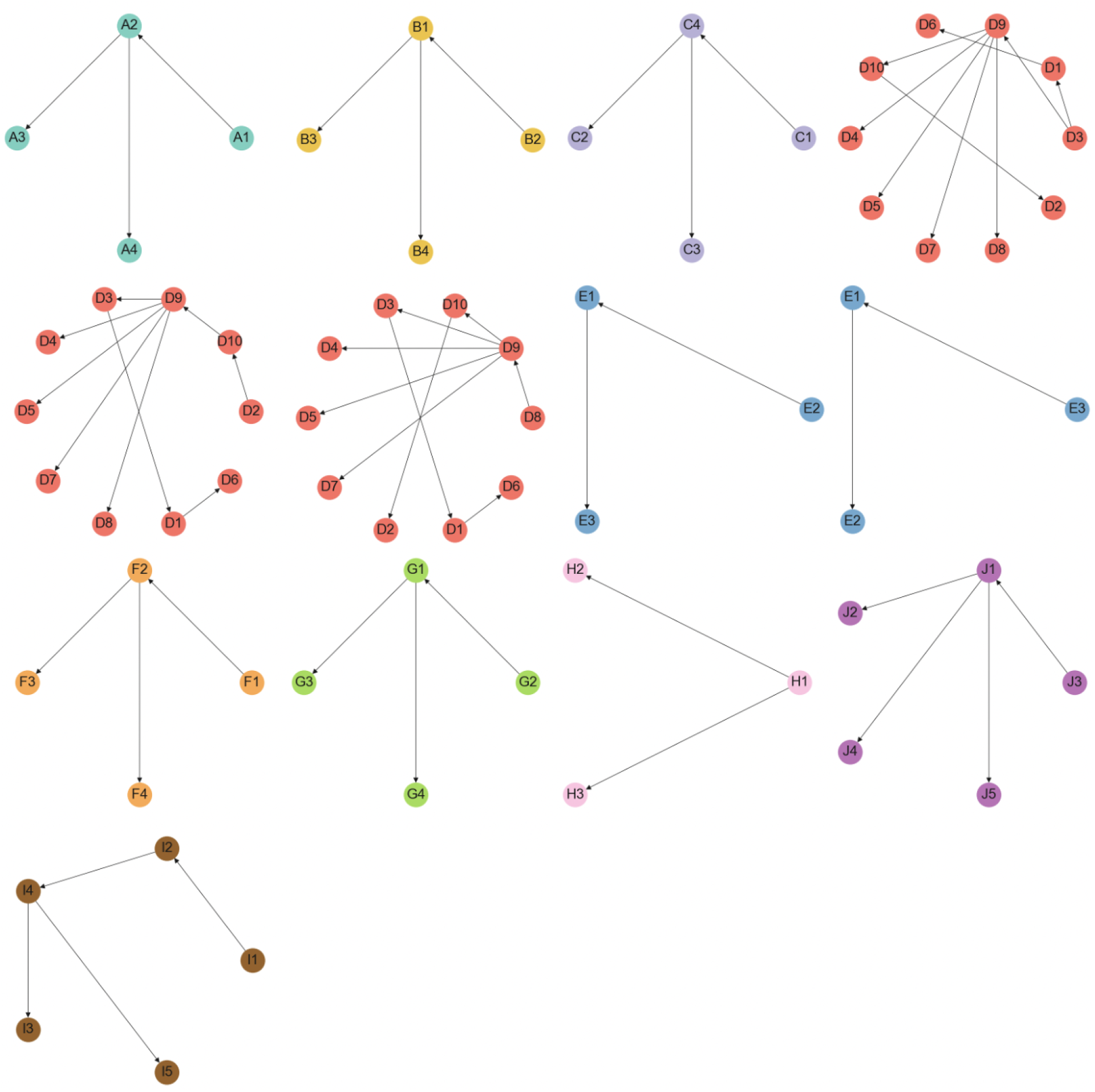
Minimum spanning trees for each cluster. Arrows indicate the inferred direction of transmission according to the arbitrary rules described in the Methods section.

**Figure S5.**
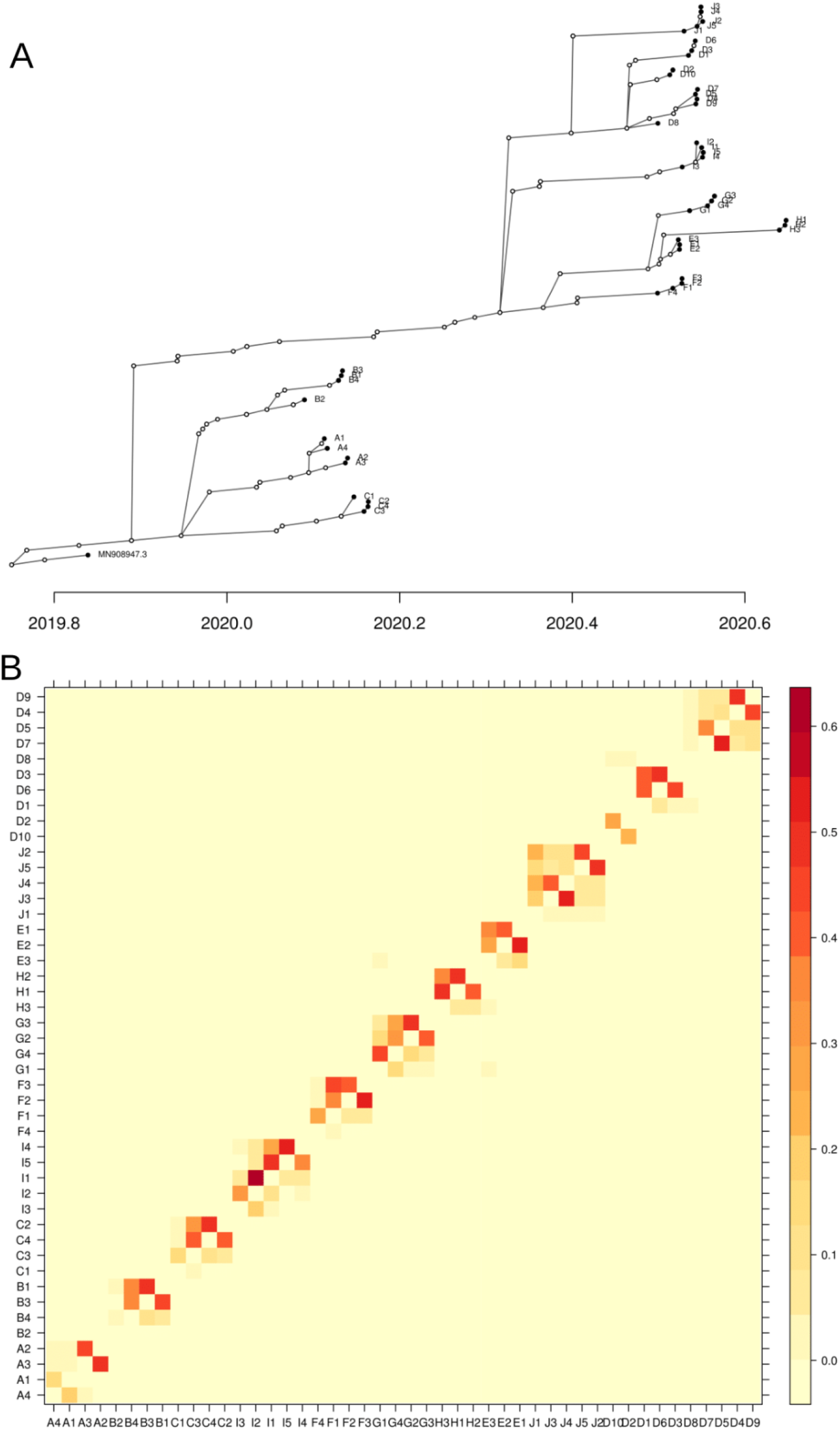
Reconstruction of transmission history using TransPhylo based on the dated phylogeny. (A) Consensus transmission tree. Filled dots represent sampled individuals, and unfilled dots represent inferred unsampled individuals. (B) Heatmap of pairwise transmission probabilities.

